# Interplay of Host and Viral Genetic Variations in Modulating Antibody Responses to Genotype 3a Hepatitis C Virus: Implications for Vaccine Design

**DOI:** 10.1101/2025.01.16.25320631

**Authors:** Zhiqing Wang, Isla Humphreys, Jocelyn Quistrebert, Haiting Chai, Josh Dhir, Alexandru Nisioi, Paul Radford, STOP-HCV consortium, Jonathan K. Ball, William L. Irving, Paul Klenerman, Eleanor Barnes, Jane A. McKeating, Alexander W. Tarr, M. Azim Ansari

**Affiliations:** Nuffield Department of Medicine, University of Oxford, Oxford OX1 3SY, UK; Institute of Infection and Immunity, The University of Birmingham; School of Life Sciences, Faculty of Medicine & Health Sciences, The University of Nottingham, Nottingham NG7 2RD, UK; Wolfson Centre for Global Virus Infections, The University of Nottingham, Nottingham NG7 2UH, UK; NIHR Biomedical Research Centre, Nottingham University Hospitals Trust, Nottingham NG7 2UH, UK; Chinese Academy of Medical Science Oxford Institute, University of Oxford, Oxford, UK; NIHR Biomedical Research Centre, Oxford University NHS trusts

**Keywords:** Hepatitis C Virus, vaccine, antibody response, neutralization, polymorphism, *IFNL4*

## Abstract

Hepatitis C virus (HCV) exhibits significant genetic diversity and is a of cause severe liver complications, including liver failure and hepatocellular carcinoma. The envelope glycoproteins E1 and E2 of HCV are the primary targets for the humoral immune response and have the highest sequence diversity in the HCV genome. The humoral immune response to HCV infection may be modulated by host and virus genetic factors. Here, using virus sequencing and host genetic data, as well as antibody binding and neutralization assays from 54 patients infected with HCV subtype 3a virus, we investigated the factors associated with antibody binding and neutralization. Genetic variation in the host *IFNL4* gene were associated with antibody binding response, with the IFNλ4-P70 variant associated with reduced binding relative to IFNλ4-Null variant. Testing for association between all variable amino acids in HCV E1 and E2 glycoproteins and antibody response, we discovered two sites in or near the CD81 binding sites in E2 (sites 501 and 533) that were associated with neutralization sensitivity, along with an additional site in E2 (653) that was associated with binding. Additionally, motifs at two glycosylation sites (N476 and N234) were associated with binding and/or neutralisation. Furthermore, an increase in intra-patient hypervariable region 1 (HVR1) diversity was associated with stronger binding response. By considering the complex factors that influence antibody binding and neutralization, future vaccine strategies can be designed to elicit a robust immune response against a diverse range of HCV strains, addressing the needs of a diverse population.

## Introduction

Elimination of hepatitis C virus as a public health concern is a World Health Organization priority to be delivered by 2030. Despite the great success of direct-acting antiviral therapies (DAA) (Cornberg & Manns, 2022), progress on developing an effective vaccine has been slow (Duncan et al., 2020). Virus-neutralizing antibodies target the HCV-encoded E1 and E2 glycoproteins that mediate viral entry (Kinchen et al., 2018; Stamataki et al., 2008). Though the molecular targets of these antibodies are well defined (Duncan et al., 2020), the antibody correlates of protection remain unclear. The E2 protein is the major receptor binding protein that directly engages with the host cell membrane entry factors (Douam et al., 2014; Dreux et al., 2009; Flint & McKeating, 2000; Liu et al., 2009; Pileri et al., 1998), while E1 protein is proposed to stabilize the structure of E2 (Torrents de la Pena et al., 2022), and contributes to receptor binding and fusion events that lead to delivery of the virus genome into a cell (Banda et al., 2019; Douam et al., 2014). The E1/E2 complex possesses a novel structure with E1 performing a ‘grasping’ mechanism to interact with the stem of E2 (Augestad et al., 2024; Torrents de la Pena et al., 2022). This structure is consistent with the E2 protein being the receptor binding protein (Kong et al., 2013), which is known to interact with the molecules CD81 and scavenger receptor class B type 1 (SR-B1) on the surface of hepatocytes (Bartosch et al., 2003; Pileri et al., 1998).

HCV is highly mutable due to the error-prone nature of genome replication mediated by the NS5B polymerase. The virus displays extensive genome plasticity (reviewed in (Echeverria et al., 2015)), with individual strains differing by more than 35% of their nucleotide sequence at the extremes of diversity. As a consequence, HCV genome sequences are classified into eight distinct genotypes. Within genotypes, many subtypes have been classified with a maximum of 20-25% difference between genomes in the same subtype. The most common HCV genotypes causing infection are 1 and 3 (Petruzziello et al., 2016) with genotype 1 being highly prevalent in Europe, America and far eastern countries and genotype 3 highly prevalent in South Asian countries (Lin et al., 2021). The genotype of the infecting virus influences the specificity of the antibody response to the E1 and E2 glycoproteins (Tarr et al., 2011), presenting a challenge for vaccine design.

Understanding the functional consequences of polymorphisms in the viral proteins informs the development of new antivirals and vaccines. Even minor differences in amino acid sequence can result in considerable functional changes. While the current generation of DAA therapies are effective against all major viral genotypes, some subtypes exhibit inherent drug resistance (Gottwein et al., 2018; Nguyen et al., 2020; Smith et al., 2019; Smith et al., 2021). HCV genotype 3 presents specific challenges, being associated with faster rates of disease progression compared to other genotypes (Chan et al., 2017) and subtype 3b is inherently more resistant to DAAs (Smith et al., 2019). Genetic variability is not uniform across the virus genome, with extreme diversity exhibited in the E1 and E2 coding genes. This has provided a barrier to development of a vaccine that can elicit antibodies capable of neutralizing the wide range of *in vivo* HCV isolates (Alzua, Pihl, Offersgaard, Duarte Hernandez, et al., 2023; Sliepen et al., 2022; Tarr et al., 2018). We have previously characterized the antibody neutralization resistance of the major HCV genotypes and subtypes (Salas et al., 2022; Tarr et al., 2011; Urbanowicz et al., 2015), identifying patterns of antibody reactivity associated with specific viral isolates. However, examining the specificity of the antibody response in polyclonal antibody preparations from natural infection is an ongoing challenge.

Glycosylation is a major and crucial co- and post-translational modification of proteins that leads to glycoprotein formation by means of various glycopeptide linkages (Spiro, 2002). The β-glycosylamine linkage of N-acetylglucosamine (GlcNAc) to asparagine (N-glycosylation) represents the most widely distributed carbohydrate–peptide bond and is the most common type of glycosylation. The binding and cleavage of the sugar-amino acid linkage are critical to the biological activity of glycoproteins. Alterations in glycosylation are associated with diverse diseases. The HCV E1 and E2 proteins are extensively glycosylated with N-linked glycans, with up to 5 sites on E1 and up to 11 sites on E2, accounting for approximately one-third of the heterodimer molecular mass (Goffard et al., 2005). Despite the high sequence heterogeneity, many of the N-glycosylation sites in the E1 and E2 proteins are conserved among the various HCV genotypes and are important for evasion of antibody binding (Alzua, Pihl, Offersgaard, Velazquez-Moctezuma, et al., 2023; Lavie et al., 2018). However, these carbohydrates provide target for interactions with the lectins that contribute to innate immune response (Brown et al., 2010; Hamed et al., 2014).

Host genetic variation is one of the factors that drives antibody responses. Genetic variations in the interferon lambda 4 (*IFNL*4) gene is associated with the spontaneous clearance and IFN-based treatment response of HCV infection (Ge et al., 2009; Thomas et al., 2009). Two polymorphisms in *IFNL4* gene, rs11322783 [ΔG > TT] and rs117648444 [G > A], create four haplotypes (Terczynska-Dyla et al., 2014), two that do not produce IFNλ4 protein (TT/G or TT/A: IFNλ4-Null) and two that express IFNλ4 protein variants (ΔG/G: IFNλ4-P70 and ΔG/A: IFNλ4-S70). Patients with the IFNλ4-Null and IFNλ4-S70 variants exhibit lower hepatic interferon-stimulated gene expression, which is associated with increased viral clearance and response to IFN-based therapy compared to those with the IFNλ4-P70 variant (Eslam et al., 2017). Additionally, genetic variation in the HLA region has been reported to be associated with spontaneous clearance of HCV infection (Huang et al., 2016).

We employed a genetic screening approach to uncover new virus polymorphisms in the HCV E1/E2 coding region linked to anti-E1/E2 antibody responses in patients with HCV genotype 3a infection. Our investigation unveiled three commonly observed amino acid substitutions in the HCV E2 protein that associated with anti-E1/E2 antibody binding and neutralization sensitivity in an HCV pseudotype-based infection model. Additionally, variants in two N-linked glycosylation sites were found to be associated with antibody responses. We also investigated the relationship between *IFNL4* gene variants and the antibody response and identified an association between the *IFNL4* haplotype related to production of IFNλ4-P70 and lower levels of antibody response. The identification of novel polymorphisms and the presence of different N-linked glycosylation motifs in the HCV E1/E2 protein associated with antibody response provide potential targets for vaccine development. In summary, our study suggests that genetic variation in both the virus and the host can impact the potency of the antibody response to HCV infection and these findings have the potential to inform the development of more effective and personalized HCV vaccines.

## Results

### Cohort description and measurement of HCV specific antibodies

Samples from 60 individuals from the BOSON clinical trial (registration number: NCT01962441) were studied (Foster et al., 2015). Utilising baseline plasma samples, the polyclonal antibody binding response directed to the full length E1 and E2 envelope glycoproteins was assessed by ELISA, using an E1/E2 expression construct produced in human HEK293T cells. As target antigen a well characterized clone (UKN3A13.6) was utilized (Owsianka et al., 2005). This expression clone was previously identified as functional in a retrovirus pseudotype infection assay, confirming expression of correctly folded protein able to utilize cellular receptors for specific entry. The sequence of this antigen clustered among our study samples in a maximum likelihood phylogenetic tree (Figure 1A) based on E1 and E2-encoding nucleotide sequences. Initial experiments titrated all sera against the reference UKN3A13.6 antigen (Supplementary Figure 1). Additionally, we used the baseline plasma samples to evaluate the neutralization sensitivity of the antibody responses in each individual using retroviruses pseudotyped with an identical full length E1/E2 construct representing UKN3A13.6. The ELISA and neutralization assays were reproducible with mean coefficient of variations of 8.5% and 14.4%, respectively. Entire consensus genome sequences for HCV were generated for 57 of the infections. Of these, 54 individuals were infected with HCV genotype 3a (gt3a), and of the remaining three, two were infected with gt3b and one with gt2b. To minimize the impact of virus genetic heterogeneity, we focused on samples from the 54 patients infected with gt3a viruses. We also examined the relationship between host *IFNL4* gene haplotypes and HLA alleles with antibody responses in the context of chronic HCV infection.

**Fig. 1.**
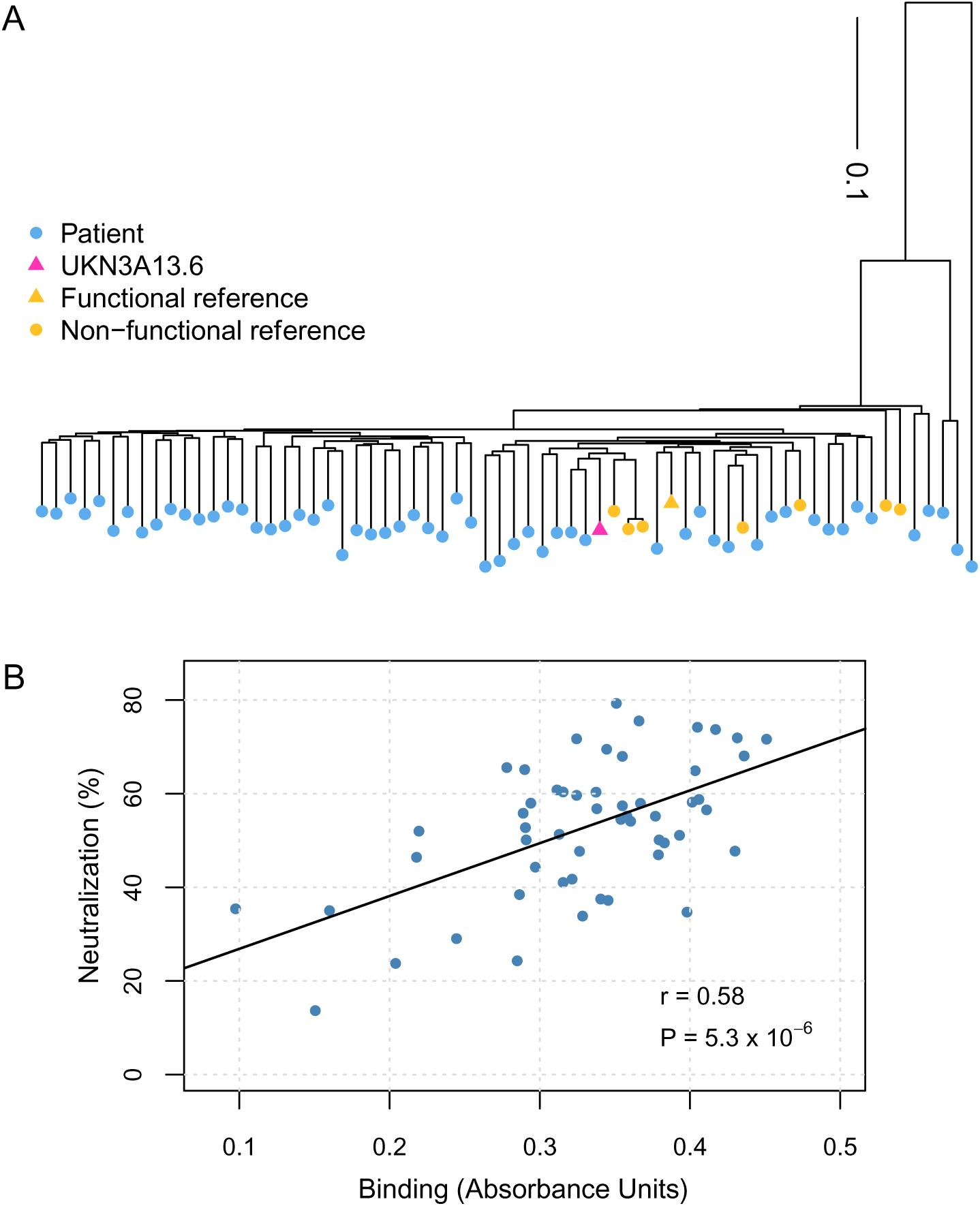
**A) Maximum likelihood phylogenetic tree showing the relationship between the study isolates (blue) and reference isolates (yellow)**. Functional and non-functional reference isolates are denoted by yellow triangles and circles respectively. The chosen reference isolate UKN3A13.6 is highlighted with a red triangle. **B) Correlation between binding and neutralization**. The solid black line represents the best fit linear regression line where the p-value for the slope being non-zero is *P =* 5.3 × 10^-6^.

After performing antibody binding and neutralization assays, we used a linear regression analysis to determine whether binding and neutralization were associated with number of amino acid differences between the reference UKN3A13.6 antigen and the HCV consensus sequences of the study samples. No significant associations between binding and number of amino acid differences (*P* = 0.36, Supplementary Figure 2A) or neutralization and number of amino acid differences (*P* = 0.4, Supplementary Figure 2B) were found. Moreover, we observed a strong association between binding and neutralization (*P* = 5.3 × 10^-6^, Figure 1B).

### Non-genetic factors associated with antibody responses

In a multivariable linear regression analysis, we investigated the association between antibody response and several factors, including sex, cirrhosis status, prior interferon treatment, age and log_10_ viral load (Figure 2A). Binding and neutralization values were standardized to have a mean of zero and standard deviation of one. Patient’s sex had the largest impact on the antibody binding response where males had lower anti-E1E2 binding levels than females (*P* = 0.011). Cirrhosis and previous interferon treatment status were also associated with binding, where patients without cirrhosis or with previous interferon treatment displaying lower antibody binding responses (*P*_cirrhosis_ = 0.029, *P*_prior-treatment_ = 0.04). Higher viral load was associated with higher binding, but this effect was not statistically significant (*P*_viral-load_ = 0.11). We also tested the association between neutralization and the same factors using linear regression (Supplementary Figure 3A). Prior interferon treatment was marginally associated with lower levels of neutralization sensitivity (*P* = 0.044). Male sex and absence of cirrhosis were associated with lower neutralization sensitivity, but the effects were not statistically significant (*P*_male_ = 0.089, *P*_prior-reatment_ = 0.1, Supplementary Figure 3A).

**Fig. 2.**
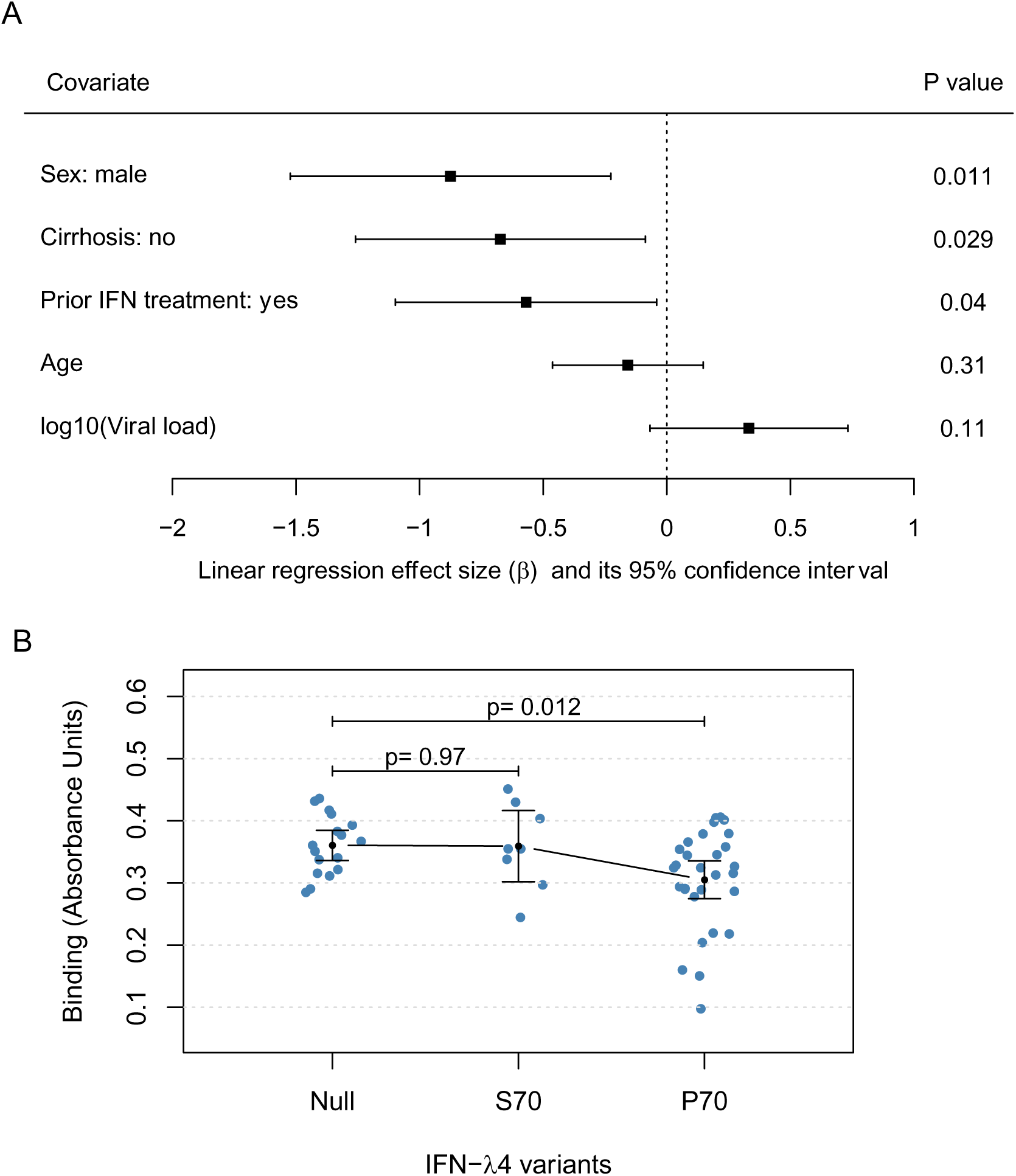
The impact of genetic and non-genetic factors on antibody binding response in HCV infection. **A) Forest plot of the effect sizes and their confidence intervals for non-genetic factors tested against binding.** The squares show the linear regression estimated effect sizes (in units of standard deviation) for each covariate and the lines show its 95% confidence interval. The *p* values for each covariate are shown on the right (n=54). **B) Binding stratified by the host IFNλ4 protein variants**. The black dots and lines indicate the mean and its 95% confidence interval for each group. The *p* values for the difference in mean relative to the IFNλ4-Null were calculated using a linear regression model (*P*_S70_ = 0.97, *P*_P70_ = 0.012).

### Host genetic factors associated with antibody responses

Since *IFNL4* gene haplotypes are highly associated with HCV spontaneous clearance and IFN-based treatment response, we hypothesized they may also be associated with antibody responses. We examined the association between the three haplotypes linked to the three protein variants (IFNλ4-Null, IFNλ4-S70 and IFNλ4-P70) and antibody responses (Figure 2B and Supplementary Figure 3B). We employed a dominant genetic model, where individuals with one or two copies of IFNλ4-P70 were classified as IFNλ4-P70, while individuals with two copies of IFNλ4-S70 or one copy of IFNλ4-S70 and one copy of IFNλ4-Null were classified as IFNλ4-S70. Patients classified as IFNλ4-P70 were observed to have significantly lower antibody binding than patients with IFNλ4-Null (*P* = 0.012) in regression analysis. However, antibody binding levels were similar in patients with IFNλ4-S70 and IFNλ4-Null variants (*P* = 0.97). We observed a modest reduction in antibody neutralization sensitivity in IFNλ4-P70 individuals compared to the other two groups, although, the result was not statistically significant (Supplementary Figure 3B). We also investigated the association between HLA alleles and antibody response in HCV infection (Supplementary Table 1 and Supplementary Table 2). HLA-A*03:01 allele was nominally associated with reduction in antibody neutralization sensitivity (*P* = 0.023), but after multiple testing correction the effect was not statistically significant.

### Virus genetic factors associated with antibody responses

We tested for associations between consensus amino acid variants in E1 and E2 proteins and antibody response using linear regression analysis. Sex, cirrhosis status, prior IFN-based treatment, and *IFNL4* haplotypes were included as covariates to minimize possible confounding effects. In our analysis, we used binding and neutralization data as response variables and the presence or absence of amino acid residues in polymorphic sites of E1 and E2 glycoproteins as explanatory variables. We only tested residues that were present in at least ten isolates which resulted in testing 123 residues (at 77 sites) in each of the assays (Supplementary Table 3 and Supplementary Table 4). At a false-discovery rate (FDR) of 20%, we found one viral polymorphism significantly associated with antibody binding (Figure 3A), and two with neutralization sensitivity (Figure 3B). Specifically, residues at site 653 in E2 protein (numbering relative to H77 polyprotein, most common sequence surrounding the site 653 RGERC**D**IEDRD) were significantly associated with antibody binding (Figure 3A), where asparagine at this site was associated with reduced binding relative to aspartic acid (Figure 3C, *P* = 6.8 × 10^-5^). However, this site was not associated with antibody neutralization (*P* = 0.19, Supplementary Figure 4A). Amino acid variations at sites 501 and 533 in E2 protein were associated with antibody neutralization sensitivity, where asparagine at site 501 was associated with higher levels of neutralization sensitivity relative to other residues (*P* = 0.001, Figure 3D, most common sequence surrounding the site 501 IVPAL**N**VCGPV). At site 533, possessing a glutamic acid residue was associated with higher neutralization sensitivity (*P* = 0.0034) compared to other residues at this site (Figure 3E, most common sequence surrounding the site 533 TWGEN**E**TDVFL). The impact of these residues on binding was consistent with their impact on neutralization sensitivity, however the effects were non-significant (*P*_501_ = 0.067, *P*_533_ = 0.053, Supplementary Figure 4B and 4C). These three sites are highly variable in wildtype gt3a isolates. To see the frequency of the amino acids at these three associated sites in the full BOSON dataset (n = 507), we calculated the frequency of all the observed amino acids as shown in the Supplementary Table 5.

**Fig. 3.**
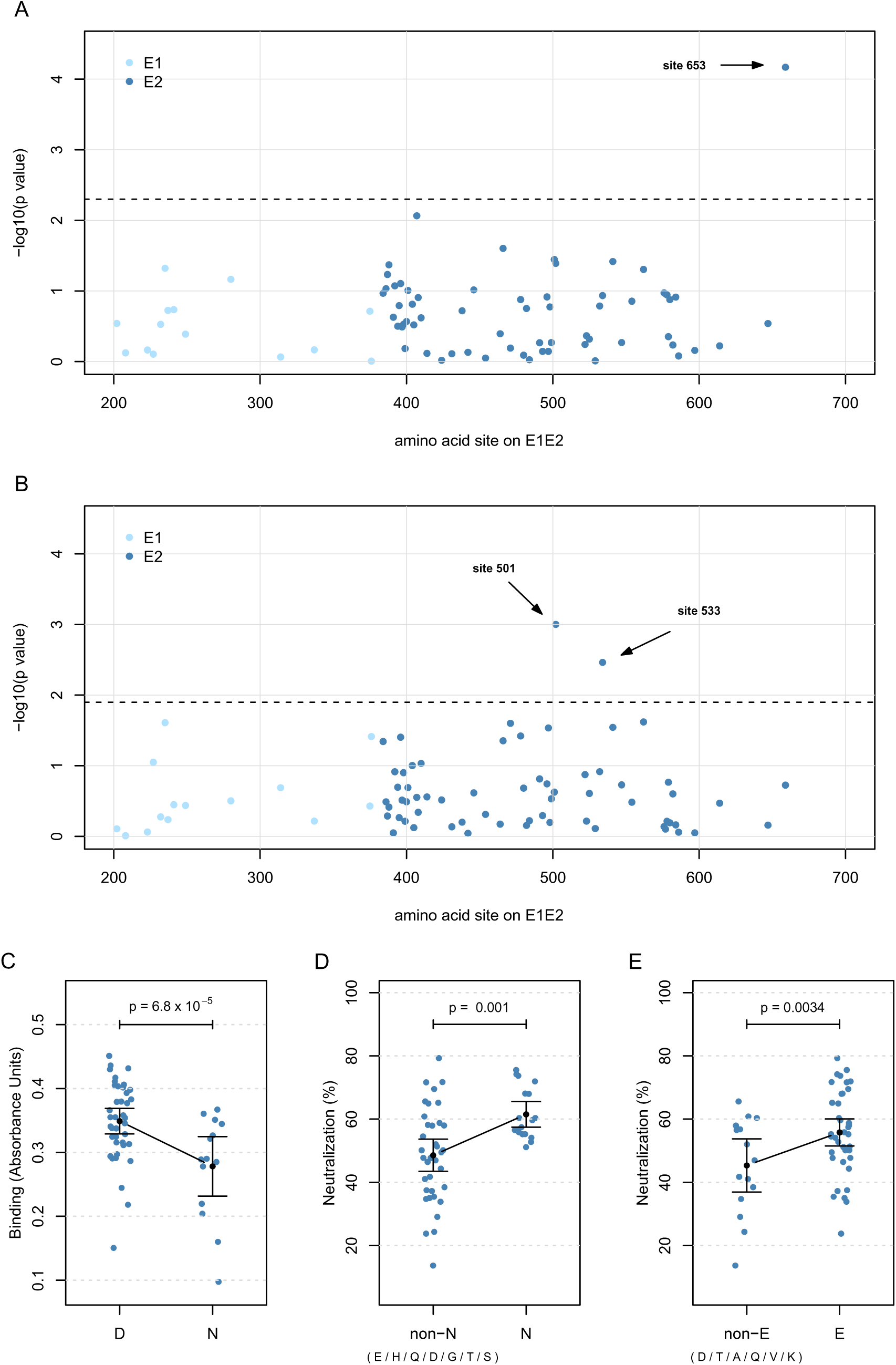
Association between HCV amino acid polymorphisms in E1 and E2 proteins and antibody response. **A) Association with binding response.** Site 653 (relative to the H77 polyprotein) is significantly associated with antibody binding response at 20% false discovery rate (FDR). **B) Association with antibody neutralization sensitivity.** Sites 501 and 533 are significantly associated with neutralization sensitivity at 20% FDR. For panels **C**, **D** and **E**, the black dots and lines indicate the mean and 95% confidence interval (CI) for each group. **C) Site 653 is associated with binding response.** Two amino acids, aspartic acid (D) and asparagine (N) were observed at site 653. *P =* 6.8 × 10^-5^ indicated the binding difference between patients carrying D and N at site 653, calculated using linear regression. **D) Site 501 is associated with neutralization sensitivity.** Asparagine (N) at site 501 was the most associated residue. *P =* 0.001 was from linear regression indicating neutralization difference between patients with N and non-N residues at site 501. The amino acids listed in the non-N group are ordered in increasing frequency. **E) Site 533 is associated with neutralization sensitivity.** Glutamic acid (E) at site 533 was the most associated residue. *P =* 0.0034 was calculated using linear regression. The amino acids listed in the non-E group are ordered in increasing frequency.

To experimentally assess the phenotypic impact of the polymorphisms associated with neutralization, we evaluated the impact of the relevant polymorphisms (T501N and E533K) on viral fitness in our HCV pseudotype model. However, both polymorphisms were associated with extremely low levels of infectivity and as such it was not possible to assess their relevance to antibody-mediated neutralization. This highlights the importance of these sites in virus for cell entry (Supplementary Figure 5).

### Virus glycosylation is associated with antibody responses

We conducted a linear regression analysis to test for association between the changes in glycosylation motifs in E1 and E2 and antibody responses. Antibody neutralisation and binding was used as the outcome variable, while the presence or absence of different glycosylation motifs at each glycosylation site was used as the exposure variable. To control the possible confounding factors, sex, cirrhosis status, prior IFN-based treatment, *IFNL4* haplotypes and log10 of baseline viral load were included as covariates in the model. Most of the sequences (48/54) carried 15 glycosylated sites in their E1E2 proteins (Supplementary Table 6). We observed six additional glycosylation sites that occurred in only one or two sequences. Of the 15 conserved glycosylated sites, six sites carried at least 2 different motifs with frequencies greater than 10. We only performed the linear regression at these six sites. We found that motifs at two glycosylation sites were associated with binding and/or neutralisation at 20% FDR. Respectively, motif NIT at site N476 (in E2 protein, numbering relative to H77 polyprotein) was significantly associated with higher levels of binding and neutralization response compared to non-NIT motifs (Figure 4A *P_binding_* = 0.035 and 4B *P_neutralisation_* = 0.0044). Additionally, the NTS motif at site N234 (in E1 Protein, numbering relative to H77 polyprotein) was found associated with reduced neutralization sensitivity, relative to non-NTS (Figure 4C, *P* = 0.021). This glycosylation site was not statistically significant in its association with binding response, however NTS motif was associated with lower binding response (Supplementary Figure 6, *P* = 0.059).

**Fig. 4.**
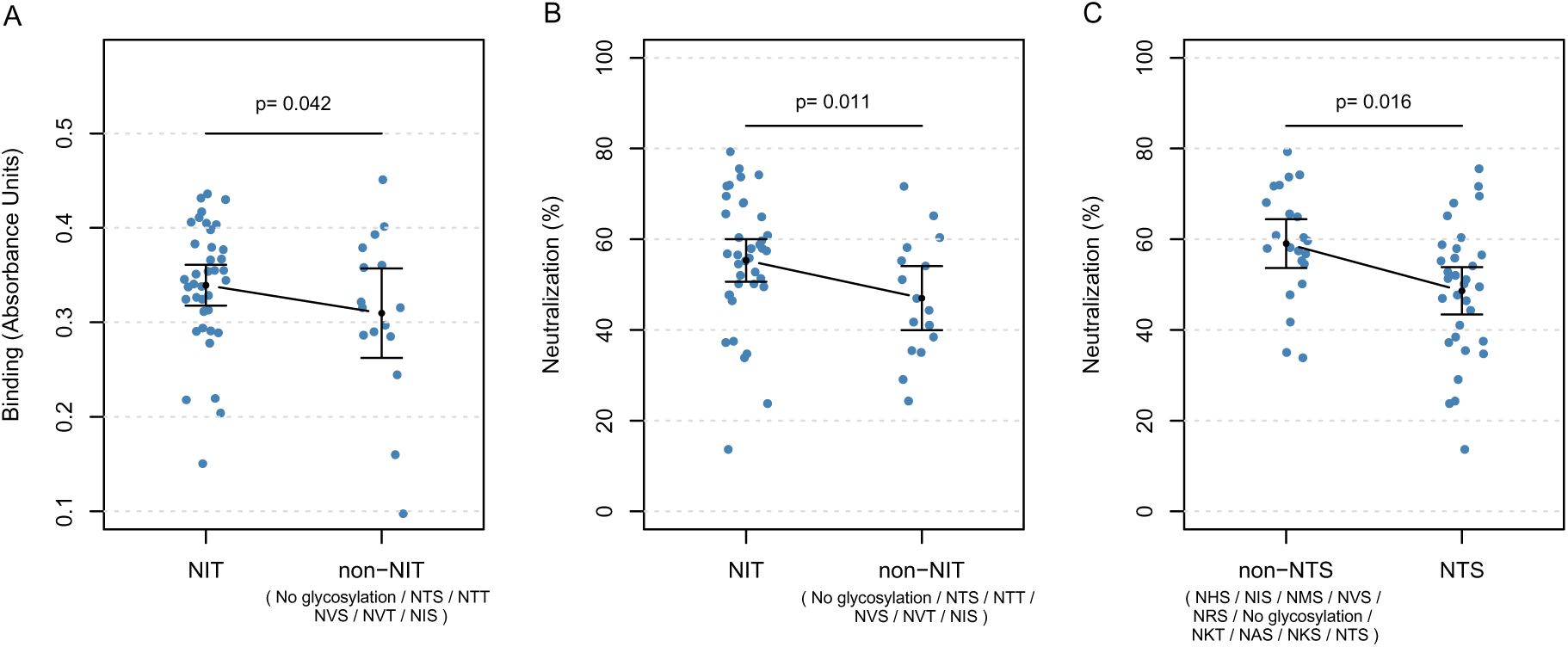
Association between variants in HCV glycosylation sites in E1 and E2 proteins and antibody response. **A) Site N476 is significantly associated with antibody binding response at 20% false discovery rate (FDR).** Motif NIT at this site was associated with higher levels of binding relative to non-NIT motifs *(P =* 0.035 from linear regression). The motifs listed in the non-NIT group are ordered in increasing frequency. **B) Site N476 is significantly associated with neutralization sensitivity at 20% FDR.** Motif ‘NIT’ at site N476 was associated with higher levels of neutralization (*P =* 0.0044 from linear regression) relative to non-NIT motifs at site N476. The motifs listed in the non-NIT group are ordered in increasing frequency. **C) Site N234 is significantly associated with neutralization sensitivity at 20% FDR.** Motif NTS at site N234 is associated with lower levels of neutralization (*P =* 0.021 from linear regression) relative to non-NTS motifs. The motifs listed in the non-NTS group are ordered in decreasing frequency. The black dots and lines indicate the mean and 95% confidence interval (CI) for each group.

### The association of intra-patient viral nucleotide diversity and antibody responses

We investigated the relationship between intra-patient viral diversity at the nucleotide level and antibody response. Nucleotide diversity, calculated from next-generation sequencing reads, was used to measure intra-patient viral diversity. We used linear regression with binding as the outcome variable and intra-patient diversity as the exposure variable to test for associations between antibody responses and the mean nucleotides diversity across the entire genome, in E1 and E2 genes (Supplementary Figure 7A), and the hypervariable region 1 (HVR1). We found a strong association between intra-patient viral diversity in HVR1 and binding response (Figure 5, *P* = 0.0016), where an increase in intra-patient HVR1 diversity was associated with greater binding. However, there were no significant association between intra-patient viral diversity and neutralization sensitivity (Supplementary Figure 7B).

**Fig. 5.**
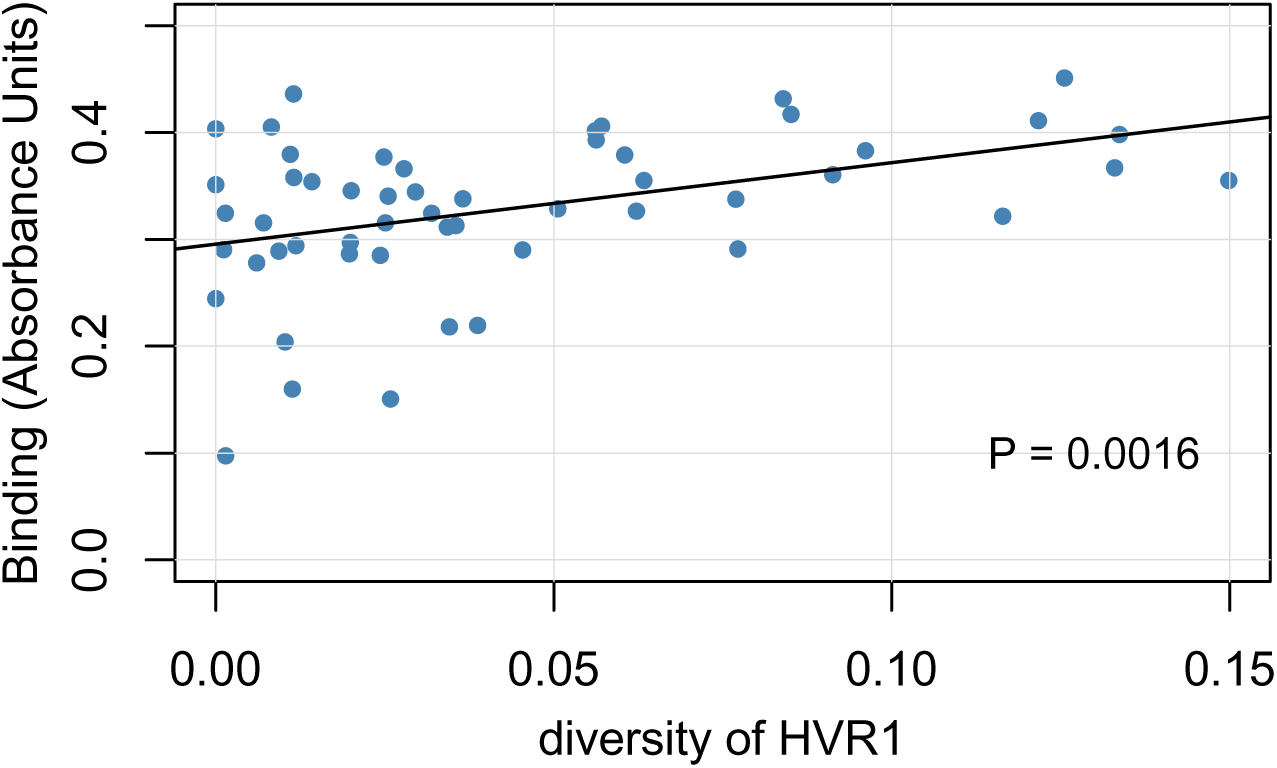
Correlation between intra-patient viral nucleotide diversity in HVR1 and antibody binding. X-axis indicates the mean nucleotide diversity in hypervariable region 1 and the y axis is the antibody binding response. The black line shows the best fit linear regression line and grey area indicates its 95% confidence interval. *P* = 0.0016 calculated using regression.

## Discussion

We report the analysis of associations between amino acid variation in HCV E1 and E2 protein sequences and host antibody responses in a well characterized patient cohort. We also examined the impact of host *IFNL4* genetic variation, HLA alleles and clinical phenotypes on anti-E1E2 responses. We discriminated between antibodies binding to E1 and E2 glycoproteins, and the neutralizing response that could limit HCV pseudo-particle infection. The specificity of the polyclonal response will influence the sensitivity of neutralization, as some antibodies that react with E1/E2 do not result in inhibition of entry (Brasher et al., 2021). The non-neutralizing antibodies may have other antiviral functions *in vivo* (Long et al., 2017), such as antibody-dependent cellular cytotoxicity (ADCC) which are not measured in our neutralization assay. We used linear regression to test for association between virus amino acid polymorphisms in E1 and E2 and antibody responses, identifying three amino acid sites in the gt3a E2 protein that impact on the antibody responses. Additionally, distinct *N*-glycan motifs present at two specific *N*-glycosylation sites in E1 and E2 were associated with the quality of antibody responses directed to these proteins. Furthermore, we found that intra-patient virus nucleotide diversity in the HVR1 region was associated with host antibody responses against E1/E2. We also observed that host *IFNL4* gene polymorphisms were associated with antibody responses as well as other factors such as sex and cirrhosis status of the patient.

Three sites in the E2 protein possessed amino acid polymorphisms associated with differential anti-E1/E2 antibody responses in gt3a virus-infected patients. Among these sites, the asparagine (N)/aspartic acid (D) residues at position 653 were found to be associated with antibody binding, which is part of a region previously reported to be targeted by non-neutralizing antibodies (Kumar et al., 2023). This polymorphism is located in a region of the E2 stem (Supplementary Figure 8) known to be immunogenic in experimental immunizations, with murine monoclonal antibodies ALP11, AP266, ALP1, ALP98, and H52 specifically targeting this region (Clayton et al., 2002). The data presented here indicates that this polymorphism influences binding of antibodies generated in natural infections. Our reference E1/E2 antigen utilized in the assays possessed an aspartic acid residue at this site, and we observed higher binding in patients whose infecting virus carried aspartic acid compared to those with asparagine at this site. One possible explanation for this difference is that the change in amino acid from D to N may result in a localized change in structure, rendering antibodies that target this region specific for one variant unable to bind the other variants. However, the drop in binding levels could also indicate that the amino acid change in this stem region results in a conformational change, causing other regions of the protein to change shape or become more, or less, accessible.

The amino acid polymorphisms observed to influence the neutralizing antibody response located at positions 501 and 533 are presented at neutralizing face of the E2 ectodomain (Tzarum et al., 2018) overlapping with the CD81 binding site. Site 533 is located in a relatively conserved region (533-535) critical for CD81 receptor interaction (Owsianka et al., 2006). The majority of viruses in our study possessed a glutamic acid (E) at this site although we observed several other residues at this site including K, V, Q, A, T and D (Supplementary Table 5). The reference antigen encoded a glutamic acid residue at site 533 and we observed higher levels of neutralization sensitivity in plasma from patients whose virus possessed a glutamic acid at this site relative to other residues. Previously it has been reported that amino acid residues in the discontinuous regions aa412-420 and aa434-446 (in E2 protein) contribute to conformational changes in the CD81 binding site (Kumar et al., 2023). As the amino acid at position 501 is proximal to the CD81 receptor binding site in the tertiary structure of E2 (Supplementary Figure 8), it is plausible that alteration from asparagine to other amino acids may change the presentation of the receptor binding site, retaining receptor binding function but altering the binding of antibodies recognising conformation-dependent epitopes. Indeed, mutants with the Q501A or S501A substitutions at this site have been implicated to influence monoclonal antibody (mAb) binding (Iacob et al., 2008; Pfaff-Kilgore et al., 2022), and the N501S mutation altered mAb binding despite having little effect on virus entry (Keck et al., 2009). Studies utilizing antibody production from human antibody libraries (Johansson et al., 2007) (Law et al., 2008; Mancini et al., 2009) and individual human B cells (Keck et al., 2013; Keck et al., 2019) have demonstrated that unlike experimental immunization, natural infection favours the production of conformation dependent anti-E2 responses. It is likely that our assays, which measure binding of both conformation-dependent and independent antibodies, may be influenced by individual, or combinations of key polymorphic sites.

When individual glycosylation sequons were analysed in the same manner as individual amino acids, it was revealed that two sites were associated with the antibody response in this patient cohort. N234 and N476 were previously implicated in resistance to neutralization, where the removal of these N-linked glycans in HCV gt1a mutants individually reduced sensitivity to polyclonal serum antibodies (Helle et al., 2007). It is plausible that the alternative sequons identified here modify the efficiency of glycosylation at these sites in naturally occurring polymorphic variants, impacting the sensitivity to neutralization. Further investigations are required to determine if differences in the patterns of glycosylation at sites 234 and 476 influence the efficiency or structure of carbohydrate modifications, as predicted for other virus glycoproteins (Kasturi et al., 1997).

We also investigated intra-patient viral diversity and observed a significant association between HVR1 nucleotide diversity and antibody binding response. The causal direction is difficult to determine as higher antibody response could result in more selective pressure on virus, encouraging it to develop escape mutations. These mutations, in turn, could lead to greater viral genetic diversity. Alternatively, higher viral sequence diversity could lead to a larger antibody response targeting multiple viruses.

In addition to the virus genetic variation, we showed that host *IFNL4* activity may shape the antibody response. Previous studies have shown that IFNλ4-P70 and IFNλ4-S70 variants of IFNλ4 protein have distinct phenotypes both *in vivo* and *in vitro* (Ansari et al., 2019; Terczynska-Dyla et al., 2014). Using the genome-wide genotyping data, we inferred haplotypes consisting of rs11322783 [ΔG>TT] and rs117648444 [G>A] variants of *IFNL4* SNPs in our cohort. Assuming a dominant effect (IFNλ4-P70 > IFNλ4-S70 > IFNλ4-Null), we observed that individuals with IFNλ4-P70 variant had significantly lower levels of antibody binding response relative to individuals with IFNλ4-S70 and IFNλ4-Null variants. We observed the same trend for neutralization sensitivity, but the effects were not statistically significant. The IFNλ4-Null variant (CC genotype at SNP rs12979860) was previously reported to associate with higher viral loads, increased spontaneous clearance, improved response to interferon treatment, and lower hepatic interferon-stimulated gene (ISG) expression. Our observation of higher anti-E1E2 antibody levels in individuals with the IFNλ4-Null variant suggests a potential mechanism: while individuals with the IFNλ4-P70 variant may experience lower viral loads due to higher hepatic ISG expression (which exert antiviral activity), IFNλ4-P70 itself may dampen the adaptive humoral immune response, potentially contributing to lower rates of spontaneous clearance and treatment response. These findings suggest a role for host *IFNL4* genetic variation in modulating the antibody response to HCV infection.

We acknowledge the limitations to our experimental approaches, particularly with the assessment of nuanced differences in the antibody binding and neutralization between different infections. Identification of amino acid signatures of antibody neutralization and binding in these samples does however highlight the power of this approach. HCVpp have been used widely to assess the breadth and potency of the serum antibody responses (Keck et al., 2009, Tarr et al., 2011, Kinchen et al., 2018), and in our laboratories provided highly reproducible inhibition between experiments. In addition, the antigens utilized for seroreactivity experiments were derived from intracellular proteins expressed in human cells. The protein preparations are likely to present a range of glycoforms with differing conformations. The use of GNA-capture ELISA assays was selected to enrich for E1E2 correctly processed protein heterodimers modified with high mannose carbohydrates, as observed on authentic particles (Guo et al., 2018). This well-established methodology generated results that broadly correlated with neutralization data, both in the current, and previous studies (Tarr et al., 2011).

In conclusion, we provide a detailed investigation of the impact of genetic variation in HCV and in the host *IFNL4* gene on humoral immune responses. We used a hypothesis-free approach and investigated all amino acid polymorphisms in HCV E1 and E2 proteins and discovered three novel sites associated with antibody responses. We also observed that changes in glycosylation motifs in two sites were associated with antibody responses. We also discovered that individuals with IFNλ4-P70 variants have a lower levels of antibody response. These observations suggest that both the host genetic background and the virus strain could drive the humoral immune response and together determine the outcome of infection and its control over time. Given that antibody responses are a critical component of most vaccine induced immunity, our results suggest that individuals with the IFNλ4-P70 variant may exhibit lower responses to an HCV vaccine, underscoring the need for tailored vaccine strategies. Moreover, our discovery that naturally occurring amino acid polymorphisms within a single HCV subtype (gt3a) impact neutralizing antibody responses emphasizes the importance of considering such variations when designing a broadly protective, pan-genotypic HCV vaccine. Such vaccines must address naturally occurring polymorphisms to enhance cross-reactivity against diverse variants and elicit robust humoral immune responses. Finally, the methodology employed in this study is broadly applicable to other viral infections, offering a framework to design vaccines that more effectively elicit immune responses by accounting for both host and viral genetic variability.

## Materials and Methods

### Patients and samples

The study was conducted in collaboration with STOP-HCV, a consortium sponsored by the Medical Research Council, United Kingdom, which contributed to study design. This study used samples from 60 patients with HCV infection selected from BOSON clinical trial (Foster et al., 2015) (registration number: NCT01962441). ELISA based IgG binding and retroviral pseudotype-based neutralization assays were implemented using samples recovered at baseline. All patients provided written informed consent before undertaking any study-related procedures. The study protocol was approved by each institution’s review board or ethics committee before study initiation.

### E1/E2 antigen and ELISA

The polyclonal antibody response directed to the HCV E1 and E2 envelope glycoproteins were detected using well characterized antigen representing genotype 3a (UKN3A13.6 (Owsianka et al., 2005), essentially as described previously (Tarr et al., 2011). Briefly, wells of Maxisorp 96-well plates were coated with *Galanthus nivalis* agglutinin (GNA) (Sigma; 1μg·mL^-1^) and blocked with PBS containing 5% BSA. Cell lysates of HEK293T cells transfected with constructs expressing the genes encoding these E1/E2 proteins were recovered 72 hours after transfection and diluted 1:5 in PBS containing 0.05% Tween (PBST) before adding to coated wells. After washing, patient plasma samples were added, diluted in PBST-5% BSA, between 1:100 and 1:10000 to titrate seroreactivity. A dilution of 1:300 was subsequently selected for further experimentation. Bound antibody was detected using anti-human Ig HRP and 3,5,3′,5′-tetramethylbenzidine (TMB) substrate was used to detect bound antibodies and the reaction was stopped after 15 minutes. The ELISA experiments were performed in duplicate and for analysis we used the mean for each sample.

### Neutralization assays

Murine Leukaemia Virus cores pseudotyped with the HCV E1/E2 genes representing isolate UKN3A13.6 were created as previously described (Urbanowicz et al., 2019), using a luciferase reporter construct (Bailey et al., 2019). Virus pseudotypes were incubated with sera (heat-inactivated prior to use at 56°C for 30 minutes) for one hour at 25°C before adding to target HuH7 cells for four hours at 37°C, in a 5% CO_2_ incubator. Following infection, media was exchanged, and cells incubated for a further 72 hours. Following cell lysis, infection was assayed using a Promega luciferase assay substrate. Luciferase activity was measured, and data were normalized to an uninhibited control and a preparation of pseudotypes created without a viral glycoprotein. Percentage values were calculated by normalizing to an uninhibited infection (100%) and the signal generated in an assay performed with pseudotypes lacking viral glycoprotein (0%). The neutralization experiments were performed in triplicate and for analysis we used the mean for each sample.

### Mutants

Single point mutants were generated using the wild-type UKN3A13.6 plasmid (accession AY894683.1) as template. Mutagenesis was performed using a Q5 Mutagenesis Kit (NEB) following the manufacturer’s protocol (primers available on request). Mutants T501N and E533K were generated and infectivity assessed using the pseudotype model described above.

### Sequencing

Sequencing was performed as described previously (Smith et al., 2021). Briefly, viral RNA was isolated from 500 μL plasma using the NucliSENS magnetic extraction system (bioMerieux). Libraries were prepared using the NEBNext Ultra Directional RNA Library Prep Kit for Illumina (New England BioLabs) with a maximum of 10 ng total RNA template. 500 ng of pooled library was enriched using the xGen Lockdown protocol from Integrated DNA Technologies (IDT) (Rapid Protocol for DNA Probe Hybridization and Target Capture Using an Illumina TruSeq or Ion Torrent Library (v1.0)) with equimolar-pooled 120-nt DNA oligonucleotide probes (IDT) followed by a 12-cycle, modified, on-bead, post-enrichment PCR re-amplification step. The cleaned post-enrichment library was normalized with the aid of qPCR and sequenced with 151-base paired-end reads on a single run of the Illumina MiSeq using v2 chemistry. De-multiplexed sequence-read pairs were trimmed of low-quality bases using QUASR (v7.0120) and of adaptor sequences using CutAdapt (v1.7.1). The remaining read pool was screened against a BLASTn database containing 165 HCV genomes, which covered its diversity both to choose an appropriate reference and to select those reads that formed a population for *de novo* assembly with Vicuna (v1.3). The assembly was finished with V-FAT v1.0 (http://www.broadinstitute.org/scientific-community/science/projects/viral-genomics/v-fat).

### Statistical methods

#### Phylogenetics

Whole-genome viral nucleotide consensus sequences for each patient in the study (n = 57) and gt3a HCV reference sequences (n = 9) were aligned using MAFFT (Katoh & Standley, 2013) with default settings. The alignment file containing only the E1 and E2 regions was uploaded to the NGPhylogeny server (https://ngphylogeny.fr/) and a maximum-likelihood tree was generated with default settings. We used R Statistical Software (Version 4.1.1) and “ape” and “phytools” packages (Paradis & Schliep, 2019; Revell, 2012) to read the newick format tree and root the tree at midpoint and plot it. We used linear regression in R (lm function) to test the impact of number of amino acid differences between the UKN3A13.6 reference and the viral consensus sequences for each patient.

#### Correlation between baseline binding and neutralization

To test for the relationship between baseline binding and neutralization, we used univariable linear regression model, calculating the *p* value and R squared. Binding was the explanatory variable and neutralization was the response variable in the model. The 95% confidence interval for the best-fit line was calculated by “predict” function from “stats” package in R.

#### Association between non-viral factors and antibody response

A multivariable linear regression model was used to test for the association between clinical phenotypes and antibody response. Standardized antibody response was used as the outcome variable and the patient’s sex, cirrhosis status, prior IFN-based treatment status, age and log of baseline viral load were included as covariates in the model. The antibody response data were standardized using “scale” function from “base” package in R. The *p* values for the associations were from linear regression model. The “metafor” package in R was used to plot the *p* values and 95% confidence intervals of the association test as a forest plot. To test the association between three *IFNL4* gene haplotypes and antibody response, the univariable linear regression was performed. The standardized antibody response was the explanatory variable and the *IFNL4* gene haplotypes were the response variable in the model. To test for HLA alleles association with antibody response, we used standardized antibody response as the explanatory variable and HLA alleles as response variables to perform univariable linear regressions. The HLA allele data were imputed as previously described (Ansari et al., 2017). The multiple tests were corrected at a false-discovery rate (FDR) of 20%. All the *p* values were from the linear regression models.

#### The association between viral amino acids polymorphisms in E1E2 and antibody response

We used linear regression to test for association between the presence and absence of each amino residue at variable sites E1 and E2 and antibody response, including patient’s sex, cirrhosis status, prior IFN-based treatment status, log of baseline viral load and *IFNL4* gene haplotypes as covariates. The threshold number for the tested residues was set to greater than ten isolates. False-discovery rate (FDR=20%) was to correct for multiple tests. The associated viral amino acids sites calculated from the tests were performed linear regression for association with antibody response, including the same confounding factors above. All the *p* values were from the linear regression models.

#### The correlation between the N-linked glycosylation motifs and antibody response

Linear regression model was used to test for association between the presence or absence of each N-linked glycosylation motif at glycosylated sites at E1 and E2 proteins. The multivariate model was including sex, cirrhosis status, prior IFN-based treatment status, log of baseline viral load and *IFNL4* gene haplotypes as covariates to control for possible confounding. We only tested sites where there was some variability in the motifs with a threshold count of 10 or more. False-discovery rate (FDR) of 20% was used to correct for multiple testing where we combined the p values generated from binding and neutralization association tests in one FDR procedure.

#### The correlation between within patient viral nucleotide diversity and antibody response

Univariable linear regression model was used to test for association between within patient viral diversity at the nucleotide level and antibody response. The mean nucleotide diversity was calculated from next-generation sequencing reads presenting as the measure of within patient viral nucleotide diversity. The mean nucleotide diversity across the whole genome, E1 combined with E2, E1, E2 and hypervariable region 1 were tested for the correlation with antibody response. All the *p* values were from the linear regression models.

## Data Availability

All data produced in the present study are available upon reasonable request to the authors

## Code availability

The R code used to generate the results and figures from the primary analyses described above are available from the authors on request.

## Acknowledgements

We thank HCV Research UK (funded by the Medical Research Foundation) for their assistance in handling and coordinating the release of samples for these analyses. This work was funded by a grant from the Medical Research Council (MRC) (MR/K01532X/1; to the STOP-HCV Consortium). This work was supported by the Chinese Academy of Medical Sciences (CAMS) Innovation Fund for Medical Science (CIFMS), China (grant 2018-I2M-2-002). M.A.A. is supported by a Sir Henry Dale Fellowship jointly funded by the Royal Society and the Wellcome Trust (220171/Z/20/Z). PK was funded by a Wellcome Trust grant(222426/Z/21/Z).

