## supplementary tables and figures for "Interplay of Host and Viral Genetic Variations in Modulating Antibody Responses to Genotype 3a Hepatitis C Virus: Implications for Vaccine Design"

#### Affiliations

### Supplementary

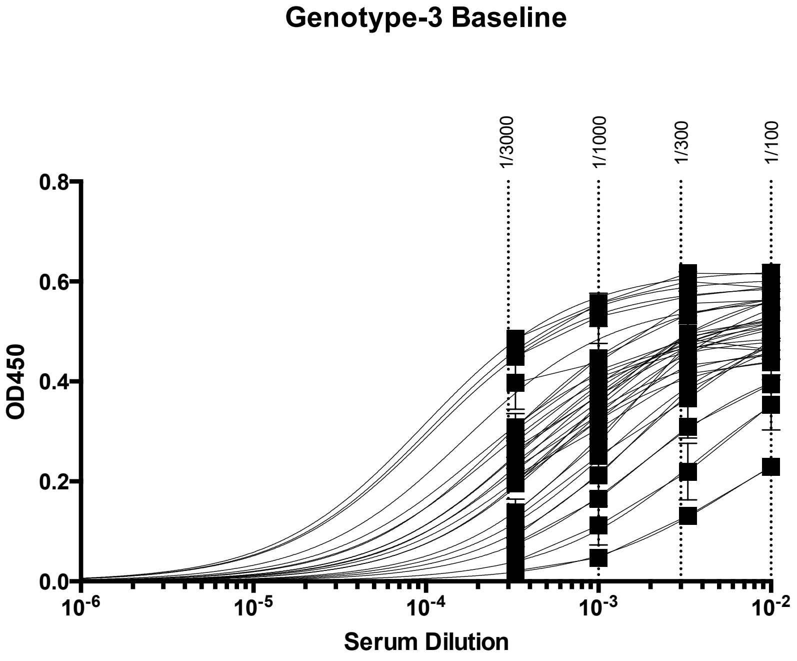


**Supplementary Figure 1. Titration of antibody reactivity to UKN3A13.6 E1/E2 proteins in ELISA**. E1/E2 proteins were expressed in HEK293T cells and antibody binding assessed using ELISA. The signal in these assays is corrected for background reactivity binding to a control cell lysate derived from mock transfected HEK293T cells. The values on the x-axis represent the dilution factor of each serum sample.

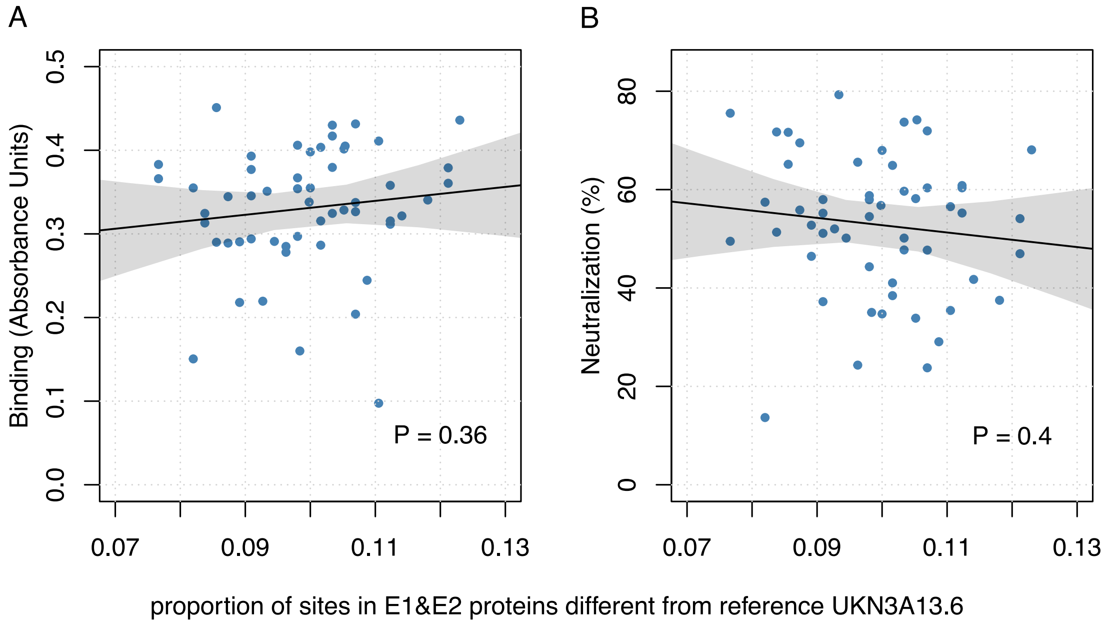


**Supplementary Figure 2: Correlation between antibody response and E1E2 amino acid divergence from the reference antigen (UKN3A13.6)**. **(A) Binding. (B) Neutralization.** Linear regression p-values for the association are shown. The blue dots indicate the proportion of differences, and the solid black lines show the best fit linear regression lines and grey area indicate its 95% confidence interval. *P*-values are estimated using a linear regression model.

A

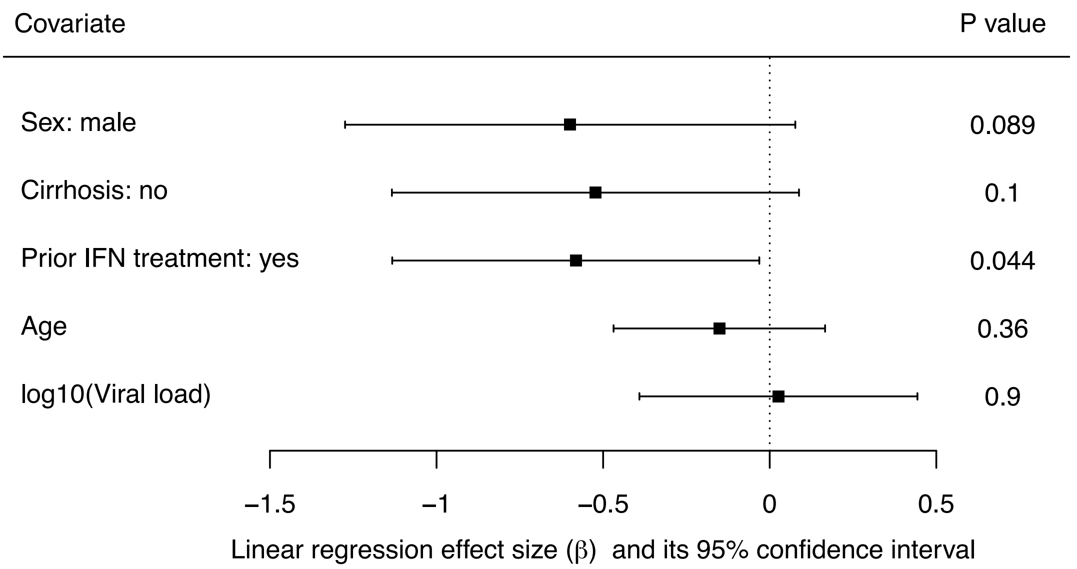


B

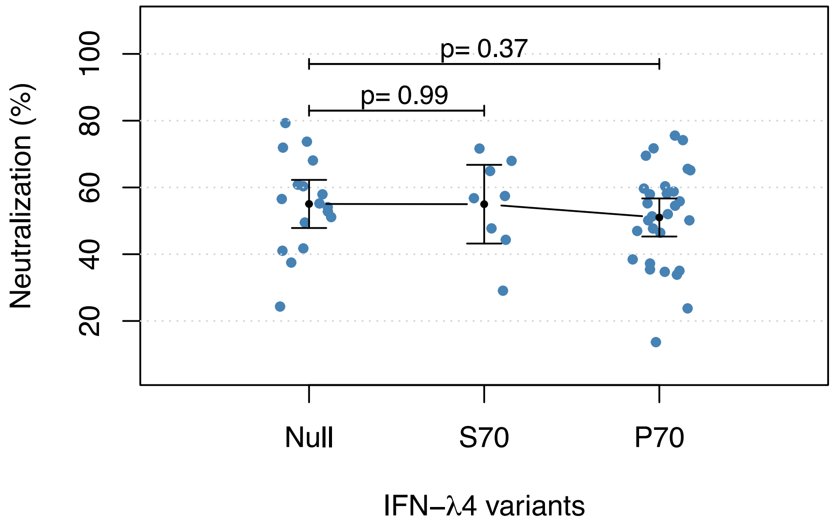


**Supplementary Figure 3: A) Forest plot of the effect sizes and their confidence intervals for non-viral factors tested against neutralization**. The squares show the linear regression estimated effect sizes for each covariate and the lines show its 95% confidence interval. The *P*-values (from linear regression) for each covariate is shown on the right (n=54). **B) Neutralization stratified by the host IFNλ4 protein haplotypes.** The black dots and lines indicate the mean and 95% confidence interval (CI) for each group. *P*-values were calculated using linear regression model.

**Supplementary Table 1. The association between HLA alleles and binding.** The tested HLA alleles (count >= 10) are listed in the table. The P values, effect size and standard error are from linear regression tests and are shown in the table. Q value was estimated using qvalue function in R.

| HLA allele | Allele count | P value | Effect size | Standard error | q value |
|---|---|---|---|---|---|
| DPA1*02:01 | 16 | 0.068 | 0.493 | 0.265 | 0.409 |
| DQB1*03:01 | 15 | 0.176 | -0.344 | 0.251 | 0.409 |
| B*07:02 | 12 | 0.195 | -0.344 | 0.262 | 0.409 |
| C*06:02 | 10 | 0.205 | -0.389 | 0.303 | 0.409 |
| DRB3*01:01 | 20 | 0.216 | -0.351 | 0.280 | 0.409 |
| C*07:02 | 14 | 0.219 | -0.317 | 0.255 | 0.409 |
| DRB3*03:01 | 12 | 0.237 | 0.314 | 0.263 | 0.409 |
| DQA1*05:01 | 21 | 0.321 | -0.241 | 0.241 | 0.409 |
| DPA1*01:03 | 51 | 0.363 | -0.209 | 0.228 | 0.409 |
| C*07:01 | 15 | 0.371 | 0.228 | 0.253 | 0.409 |
| DQB1*02:01 | 10 | 0.378 | 0.312 | 0.351 | 0.409 |
| DRB1*03:01 | 10 | 0.378 | 0.312 | 0.351 | 0.409 |
| A*03:01 | 15 | 0.391 | -0.219 | 0.253 | 0.409 |
| A*01:01 | 14 | 0.533 | 0.161 | 0.257 | 0.409 |
| DRB3*99:01 | 28 | 0.537 | 0.122 | 0.196 | 0.409 |
| A*24:02 | 10 | 0.550 | 0.212 | 0.352 | 0.409 |
| A*02:01 | 21 | 0.569 | -0.139 | 0.243 | 0.409 |
| DRB4*99:01 | 49 | 0.574 | -0.118 | 0.208 | 0.409 |
| DRB4*01:01 | 25 | 0.574 | 0.118 | 0.208 | 0.409 |
| DRB3*02:02 | 26 | 0.621 | -0.094 | 0.188 | 0.409 |
| DQA1*01:01 | 15 | 0.627 | -0.135 | 0.277 | 0.409 |
| DQA1*03:01 | 12 | 0.688 | 0.118 | 0.293 | 0.409 |
| C*04:01 | 12 | 0.700 | -0.103 | 0.266 | 0.409 |
| DQB1*03:02 | 10 | 0.701 | 0.136 | 0.353 | 0.409 |
| DRB5*99:01 | 53 | 0.787 | 0.073 | 0.270 | 0.409 |
| DQB1*06:02 | 17 | 0.787 | -0.073 | 0.270 | 0.409 |
| DRB1*15:01 | 17 | 0.787 | -0.073 | 0.270 | 0.409 |
| DRB5*01:01 | 17 | 0.787 | -0.073 | 0.270 | 0.409 |
| DQA1*02:01 | 18 | 0.806 | 0.066 | 0.267 | 0.409 |
| DRB1*07:01 | 18 | 0.806 | 0.066 | 0.267 | 0.409 |
| DQA1*01:02 | 21 | 0.827 | 0.051 | 0.229 | 0.409 |
| B*08:01 | 12 | 0.841 | 0.059 | 0.293 | 0.409 |
| DQB1*02:02 | 16 | 0.992 | 0.003 | 0.273 | 0.467 |

**Supplementary Table 2. The association between HLA alleles and neutralization.** The tested HLA alleles (count >= 10) are listed in the table. The P values, effect size and standard error are from linear regression and are shown in the table. Q value was estimated using qvalue function in R.

| HLA allele | Allele count | P value | Effect size | Standard error | q value |
|---|---|---|---|---|---|
| A*03:01 | 15 | 0.022 | -0.571 | 0.243 | 0.742 |
| DRB3*02:02 | 26 | 0.128 | -0.285 | 0.185 | 0.837 |
| DRB3*03:01 | 12 | 0.138 | 0.392 | 0.261 | 0.837 |
| A*01:01 | 14 | 0.144 | 0.375 | 0.253 | 0.837 |
| C*07:01 | 15 | 0.146 | 0.369 | 0.250 | 0.837 |
| DQA1*05:01 | 21 | 0.195 | -0.315 | 0.239 | 0.837 |
| C*07:02 | 14 | 0.215 | -0.319 | 0.255 | 0.837 |
| B*07:02 | 12 | 0.276 | -0.290 | 0.263 | 0.837 |
| A*24:02 | 10 | 0.311 | 0.358 | 0.350 | 0.837 |
| DQB1*03:01 | 15 | 0.341 | -0.243 | 0.253 | 0.837 |
| A*02:01 | 21 | 0.342 | 0.232 | 0.241 | 0.837 |
| B*08:01 | 12 | 0.383 | 0.256 | 0.291 | 0.837 |
| DQA1*01:01 | 15 | 0.450 | -0.210 | 0.276 | 0.837 |
| C*06:02 | 10 | 0.476 | -0.220 | 0.306 | 0.837 |
| DRB4*01:01 | 25 | 0.530 | 0.132 | 0.208 | 0.837 |
| DRB4*99:01 | 49 | 0.530 | -0.132 | 0.208 | 0.837 |
| DRB3*99:01 | 28 | 0.534 | 0.123 | 0.196 | 0.837 |
| DPA1*02:01 | 16 | 0.548 | 0.165 | 0.272 | 0.837 |
| C*04:01 | 12 | 0.553 | 0.159 | 0.265 | 0.837 |
| DQA1*02:01 | 18 | 0.591 | 0.144 | 0.266 | 0.837 |
| DRB1*07:01 | 18 | 0.591 | 0.144 | 0.266 | 0.837 |
| DQB1*03:02 | 10 | 0.630 | 0.171 | 0.353 | 0.837 |
| DRB3*01:01 | 20 | 0.634 | -0.136 | 0.284 | 0.837 |
| DQA1*01:02 | 21 | 0.669 | 0.098 | 0.229 | 0.837 |
| DQA1*03:01 | 12 | 0.738 | 0.098 | 0.293 | 0.837 |
| DRB5*99:01 | 53 | 0.739 | 0.090 | 0.270 | 0.837 |
| DQB1*06:02 | 17 | 0.739 | -0.090 | 0.270 | 0.837 |
| DRB1*15:01 | 17 | 0.739 | -0.090 | 0.270 | 0.837 |
| DRB5*01:01 | 17 | 0.739 | -0.090 | 0.270 | 0.837 |
| DQB1*02:02 | 16 | 0.761 | 0.084 | 0.273 | 0.837 |
| DPA1*01:03 | 51 | 0.895 | -0.030 | 0.230 | 0.897 |
| DQB1*02:01 | 10 | 0.897 | 0.046 | 0.354 | 0.897 |
| DRB1*03:01 | 10 | 0.897 | 0.046 | 0.354 | 0.897 |

**Supplementary Table 3. Statistical analysis of the association between E1/E2 amino acid variations and antibody binding.** The table presents results from linear regression models testing the association between antibody binding and polymorphisms at 77 sites (in total 123 residues were tested at these 77 sites, but only the most associated residue per site is shown in the table, numbered according to the H77 reference sequence). P-values, effect sizes, and standard errors are reported, along with q-values representing FDR-corrected p-values to account for multiple testing. Associations at 20% FDR are highlighted in bold.

| The tested site | The most associated amino acid | Amino acids in reducing frequency order at the tested site | P value | Effect size | Standard error | q value |
|---|---|---|---|---|---|---|
| **653** | **D** | **DN** | **6.80E-05** | **1.133** | **0.259** | **0.006** |
| 407 | A | APS | 0.009 | -0.698 | 0.254 | 0.372 |
| 466 | R | KRNESG | 0.025 | 0.571 | 0.246 | 0.372 |
| 500 | S | SALRTKV | 0.036 | -0.536 | 0.248 | 0.372 |
| 540 | E | EKTQNR | 0.038 | -0.553 | 0.259 | 0.372 |
| 501 | S | SNTDGEHQ | 0.041 | -0.556 | 0.264 | 0.372 |
| 388 | T | TISV | 0.043 | -0.595 | 0.285 | 0.372 |
| 235 | T | TKAHIMRV | 0.048 | -0.521 | 0.256 | 0.372 |
| 561 | V | VLTI | 0.05 | 0.489 | 0.242 | 0.372 |
| 387 | I | TVIL | 0.058 | 0.611 | 0.315 | 0.372 |
| 280 | M | MVLI | 0.068 | -0.553 | 0.297 | 0.372 |
| 396 | T | ATVILP | 0.078 | 0.611 | 0.339 | 0.372 |
| 392 | A | AVPTMQEI | 0.085 | -0.447 | 0.254 | 0.372 |
| 386 | R | YRHT | 0.093 | 0.446 | 0.26 | 0.372 |
| 446 | K | KRSQ | 0.096 | -0.468 | 0.276 | 0.372 |
| 401 | S | SGKNTAQR | 0.098 | -0.421 | 0.249 | 0.372 |
| 574 | E | GEDRAKPST | 0.106 | -0.483 | 0.293 | 0.372 |
| 384 | S | SETNQDGAHRY | 0.108 | 0.497 | 0.303 | 0.372 |
| 575 | G | GEKRMQS | 0.11 | 0.455 | 0.279 | 0.372 |
| 576 | N | NDSGEKTACPV | 0.114 | 0.475 | 0.294 | 0.372 |
| 533 | E | EKAQVDT | 0.116 | 0.457 | 0.285 | 0.372 |
| 495 | D | DGESTKNAQR | 0.121 | 0.497 | 0.315 | 0.372 |
| 578 | D | DGHNRCPS | 0.122 | 0.473 | 0.301 | 0.372 |
| 408 | Q | QKRNS | 0.124 | -0.414 | 0.264 | 0.372 |
| 478 | S | TSN | 0.132 | -0.455 | 0.297 | 0.372 |
| 576b | R | RKSNGQDHTYEP | 0.133 | -0.523 | 0.342 | 0.372 |
| 553 | T | TAVS | 0.14 | 0.377 | 0.251 | 0.372 |
| 404 | S | SNTAQKHR | 0.154 | -0.373 | 0.258 | 0.387 |

| The tested site | The most associated amino acid | Amino acids in reducing frequency order at the tested site | P value | Effect size | Standard error | q value |
|---|---|---|---|---|---|---|
| 395 | G | GSTANDQFHK | 0.161 | -0.394 | 0.276 | 0.387 |
| 531 | A | EAGTM | 0.163 | -0.38 | 0.268 | 0.387 |
| 497 | V | VI | 0.168 | -0.419 | 0.299 | 0.387 |
| 481 | D | DENAGH | 0.177 | -0.373 | 0.272 | 0.387 |
| 241 | P | PSA | 0.184 | 0.349 | 0.258 | 0.387 |
| 237 | K | TKMES | 0.188 | 0.451 | 0.338 | 0.387 |
| 438 | I | ILMV | 0.191 | -0.391 | 0.295 | 0.387 |
| 375 | M | MIVL | 0.194 | -0.358 | 0.272 | 0.387 |
| 391 | A | SATNLQR | 0.235 | -0.391 | 0.325 | 0.437 |
| 410 | N | NKRHPS | 0.24 | -0.308 | 0.258 | 0.437 |
| 400 | A | ATVSYGKL | 0.27 | 0.28 | 0.251 | 0.466 |
| 202 | V | VI | 0.288 | -0.282 | 0.263 | 0.466 |
| 641 | D | TDSENA | 0.289 | 0.344 | 0.321 | 0.466 |
| 398 | G | GSTRVFIKM | 0.297 | -0.261 | 0.247 | 0.466 |
| 232 | D | DNTHAEQS | 0.297 | -0.278 | 0.264 | 0.466 |
| 405 | P | PLQRVMAKSTW | 0.302 | -0.277 | 0.266 | 0.466 |
| 394 | R | RHQYSFGKV | 0.315 | -0.27 | 0.266 | 0.478 |
| 397 | S | SRHNFLQYGKAEMW | 0.323 | 0.268 | 0.268 | 0.481 |
| 464 | F | FSHAYN | 0.404 | 0.215 | 0.255 | 0.544 |
| 249 | R | KRE | 0.409 | -0.214 | 0.256 | 0.544 |
| 522 | K | KREGMQ | 0.431 | 0.209 | 0.263 | 0.549 |
| 576a | P | PHRLSTFAENV | 0.443 | -0.205 | 0.265 | 0.549 |
| 524 | V | AVTM | 0.48 | 0.189 | 0.265 | 0.578 |
| 546 | S | SNGRAKQ | 0.537 | 0.169 | 0.272 | 0.609 |
| 498 | P | PQSLNAKR | 0.538 | 0.173 | 0.279 | 0.609 |
| 490 | A | AP | 0.54 | -0.186 | 0.301 | 0.609 |
| 521 | A | ARVDEISLT | 0.572 | -0.156 | 0.273 | 0.616 |
| 576d | E | EGDKTANQS | 0.582 | 0.143 | 0.259 | 0.616 |
| 608 | M | MLI | 0.599 | 0.165 | 0.311 | 0.627 |
| 471 | S | PST | 0.643 | -0.137 | 0.294 | 0.645 |
| 399 | L | FLISV | 0.655 | 0.128 | 0.286 | 0.645 |
| 337 | V | VIL | 0.683 | 0.131 | 0.319 | 0.645 |
| 223 | A | TAI | 0.686 | -0.129 | 0.317 | 0.645 |
| 591 | E | EDGAK | 0.694 | 0.12 | 0.302 | 0.645 |
| 492 | R | RK | 0.716 | -0.107 | 0.294 | 0.645 |
| 496 | V | TIVDELNS | 0.717 | 0.118 | 0.323 | 0.645 |

| The tested site | The most associated amino acid | Amino acids in reducing frequency order at the tested site | P value | Effect size | Standard error | q value |
|---|---|---|---|---|---|---|
| 442 | F | FIVL | 0.739 | 0.106 | 0.316 | 0.645 |
| 208 | S | SP | 0.752 | 0.102 | 0.321 | 0.645 |
| 414 | V | VI | 0.764 | -0.078 | 0.258 | 0.645 |
| 431 | D | DEA | 0.775 | -0.076 | 0.265 | 0.645 |
| 227 | I | IV | 0.785 | 0.076 | 0.278 | 0.645 |
| 479a | S | PSDT | 0.813 | -0.066 | 0.277 | 0.661 |
| 580 | F | FLIHVAMSTY | 0.831 | 0.059 | 0.275 | 0.663 |
| 314 | S | ST | 0.861 | -0.048 | 0.274 | 0.663 |
| 454 | Q | QEHRDGLY | 0.892 | -0.044 | 0.323 | 0.681 |
| 483 | K | KR | 0.942 | 0.02 | 0.269 | 0.705 |
| 424 | R | RS | 0.959 | -0.014 | 0.272 | 0.705 |
| 528 | T | TNSDGQ | 0.977 | 0.009 | 0.302 | 0.705 |
| 376 | V | VI | 0.985 | -0.006 | 0.315 | 0.705 |

**Supplementary Table 4. Statistical analysis of the association between E1/E2 amino acid variations and antibody neutralization.** The table presents results from linear regression models testing the association between antibody binding and polymorphisms at 77 sites (in total 123 residues were tested at these 77 sites, but only the most associated residue per site is shown in the table, numbered according to the H77 reference sequence). P-values, effect sizes, and standard errors are reported, along with q-values representing FDR-corrected p-values to account for multiple testing. Associations at 20% FDR are highlighted in bold.

| The tested site | The most associated amino acid | Amino acids in reducing frequency order at the tested site | P value | Effect size | Standard error | q value |
|---|---|---|---|---|---|---|
| **501** | **N** | **SNTDGEHQ** | **0.001** | **0.963** | **0.274** | **0.108** |
| **533** | **E** | **EKAQVDT** | **0.003** | **0.909** | **0.295** | **0.187** |
| 561 | V | VLTI | 0.024 | 0.616 | 0.264 | 0.403 |
| 235 | T | TKAHIMRV | 0.025 | -0.649 | 0.279 | 0.403 |
| 471 | P | PST | 0.025 | 0.691 | 0.298 | 0.403 |
| 540 | E | EKTQNR | 0.029 | -0.644 | 0.285 | 0.403 |
| 496 | V | TIVDELNS | 0.029 | 0.765 | 0.34 | 0.403 |
| 478 | S | TSN | 0.038 | -0.686 | 0.321 | 0.403 |
| 376 | V | VI | 0.039 | 0.708 | 0.333 | 0.403 |
| 396 | V | ATVILP | 0.039 | -0.785 | 0.37 | 0.403 |
| 466 | K | KRNESG | 0.044 | -0.545 | 0.263 | 0.403 |
| 384 | S | SETNQDGAHRY | 0.045 | 0.678 | 0.33 | 0.403 |
| 227 | I | IV | 0.089 | -0.516 | 0.298 | 0.568 |
| 410 | K | NKRHPS | 0.093 | 0.526 | 0.306 | 0.568 |
| 404 | S | SNTAQKHR | 0.099 | -0.475 | 0.283 | 0.568 |
| 531 | E | EAGTM | 0.121 | 0.442 | 0.28 | 0.618 |
| 392 | A | AVPTMQEI | 0.122 | -0.444 | 0.282 | 0.618 |
| 398 | G | GSTRVFIKM | 0.125 | -0.42 | 0.269 | 0.618 |
| 521 | A | ARVDEISLT | 0.134 | -0.452 | 0.296 | 0.63 |
| 490 | A | AP | 0.154 | -0.474 | 0.327 | 0.663 |
| 576a | P | PHRLSTFAENV | 0.172 | 0.393 | 0.283 | 0.663 |
| 495 | D | DGESTKNAQR | 0.181 | 0.476 | 0.35 | 0.663 |
| 546 | S | SNGRAKQ | 0.187 | 0.397 | 0.296 | 0.663 |
| 653 | D | DN | 0.189 | 0.445 | 0.334 | 0.663 |
| 394 | H | RHQYSFGKV | 0.202 | -0.45 | 0.347 | 0.663 |
| 401 | G | SGKNTAQR | 0.204 | -0.406 | 0.315 | 0.663 |
| 314 | S | ST | 0.205 | 0.383 | 0.298 | 0.663 |

| The tested site | The most associated amino acid | Amino acids in reducing frequency order at the tested site | P value | Effect size | Standard error | q value |
|---|---|---|---|---|---|---|
| 479a | S | PSDT | 0.208 | -0.385 | 0.301 | 0.663 |
| 500 | S | SALRTKV | 0.236 | -0.34 | 0.283 | 0.693 |
| 446 | K | KRSQ | 0.242 | -0.366 | 0.309 | 0.693 |
| 524 | V | AVTM | 0.247 | 0.341 | 0.29 | 0.693 |
| 576d | G | EGDKTANQS | 0.251 | 0.366 | 0.314 | 0.693 |
| 414 | V | VI | 0.276 | 0.31 | 0.282 | 0.693 |
| 407 | P | APS | 0.281 | -0.344 | 0.315 | 0.693 |
| 498 | P | PQSLNAKR | 0.294 | 0.325 | 0.306 | 0.693 |
| 424 | R | RS | 0.306 | -0.308 | 0.297 | 0.693 |
| 397 | S | SRHNFLQYGKAEMW | 0.307 | -0.306 | 0.296 | 0.693 |
| 280 | M | MVLI | 0.315 | -0.341 | 0.336 | 0.693 |
| 400 | A | ATVSYGKL | 0.324 | -0.278 | 0.278 | 0.693 |
| 386 | Y | YRHT | 0.325 | 0.276 | 0.277 | 0.693 |
| 553 | T | TAVS | 0.328 | 0.277 | 0.281 | 0.693 |
| 608 | L | MLI | 0.339 | -0.349 | 0.361 | 0.693 |
| 241 | S | PSA | 0.356 | -0.298 | 0.319 | 0.693 |
| 249 | K | KRE | 0.366 | -0.263 | 0.288 | 0.696 |
| 375 | M | MIVL | 0.373 | 0.273 | 0.303 | 0.697 |
| 388 | T | TISV | 0.386 | -0.286 | 0.327 | 0.698 |
| 408 | K | QKRNS | 0.457 | -0.239 | 0.318 | 0.774 |
| 454 | Q | QEHRDGLY | 0.489 | 0.248 | 0.356 | 0.804 |
| 492 | R | RK | 0.509 | 0.215 | 0.323 | 0.815 |
| 387 | I | TVIL | 0.515 | -0.236 | 0.36 | 0.815 |
| 232 | D | DNTHAEQS | 0.531 | -0.185 | 0.294 | 0.815 |
| 395 | G | GSTANDQFHK | 0.543 | 0.19 | 0.311 | 0.815 |
| 237 | T | TKMES | 0.58 | -0.173 | 0.311 | 0.815 |
| 483 | K | KR | 0.602 | -0.156 | 0.297 | 0.815 |
| 337 | I | VIL | 0.609 | 0.187 | 0.362 | 0.815 |
| 522 | K | KREGMQ | 0.609 | 0.15 | 0.292 | 0.815 |
| 399 | F | FLISV | 0.612 | -0.154 | 0.301 | 0.815 |
| 576 | D | NDSGEKTACPV | 0.612 | 0.164 | 0.321 | 0.815 |
| 438 | I | ILMV | 0.631 | -0.16 | 0.331 | 0.815 |
| 497 | V | VI | 0.636 | -0.161 | 0.337 | 0.815 |
| 576b | R | RKSNGQDHTYEP | 0.643 | 0.179 | 0.382 | 0.815 |
| 464 | F | FSHAYN | 0.673 | 0.12 | 0.283 | 0.815 |
| 578 | D | DGHNRCPS | 0.689 | -0.137 | 0.34 | 0.815 |

| The tested site | The most associated amino acid | Amino acids in reducing frequency order at the tested site | P value | Effect size | Standard error | q value |
|---|---|---|---|---|---|---|
| 641 | T | TDSENA | 0.694 | -0.117 | 0.296 | 0.815 |
| 481 | D | DENAGH | 0.701 | -0.118 | 0.306 | 0.815 |
| 574 | G | GEDRAKPST | 0.728 | 0.102 | 0.292 | 0.829 |
| 431 | D | DEA | 0.734 | -0.1 | 0.293 | 0.829 |
| 405 | P | PLQRVMAKSTW | 0.754 | 0.094 | 0.297 | 0.835 |
| 528 | T | TNSDGQ | 0.775 | -0.096 | 0.333 | 0.841 |
| 202 | V | VI | 0.782 | -0.082 | 0.294 | 0.841 |
| 575 | G | GEKRMQS | 0.791 | 0.088 | 0.33 | 0.841 |
| 223 | T | TAI | 0.869 | -0.056 | 0.34 | 0.851 |
| 580 | F | FLIHVAMSTY | 0.875 | -0.048 | 0.304 | 0.851 |
| 591 | E | EDGAK | 0.893 | -0.045 | 0.335 | 0.851 |
| 391 | A | SATNLQR | 0.894 | 0.049 | 0.365 | 0.851 |
| 442 | F | FIVL | 0.908 | -0.04 | 0.349 | 0.857 |
| 208 | S | SP | 0.98 | -0.009 | 0.356 | 0.876 |

**Supplementary Table 5. The frequency of different amino acids at each site (site 653, 501, 533) using 507 gt3a isolates in the BOSON cohort.** The top, medium and bottom tables indicate the frequency of different amino acids at site 653, site 501 and at site 533 respectively in the BOSON dataset. The counts and percent for each amino acid in 507 gt3a samples from BOSON cohort are shown in the tables.

| Amino acid at site 653 | Count | Percent |
|---|---|---|
| D | 406 | 80.08% |
| N | 90 | 17.75% |
| E | 10 | 1.97% |

| Amino acid at site 501 | Count | Percent |
|---|---|---|
| S | 181 | 35.70% |
| N | 159 | 31.36% |
| T | 59 | 11.64% |
| D | 30 | 5.92% |
| G | 19 | 3.75% |
| K | 19 | 3.75% |
| E | 17 | 3.35% |
| R | 16 | 3.16% |
| Q | 4 | 0.79% |
| H | 1 | 0.20% |

| Amino acid at site 533 | Count | Percent |
|---|---|---|
| E | 402 | 79.29% |
| K | 52 | 10.26% |
| A | 14 | 2.76% |
| Q | 13 | 2.56% |
| D | 11 | 2.17% |
| V | 8 | 1.58% |
| T | 2 | 0.39% |
| N | 1 | 0.20% |
| P | 1 | 0.20% |

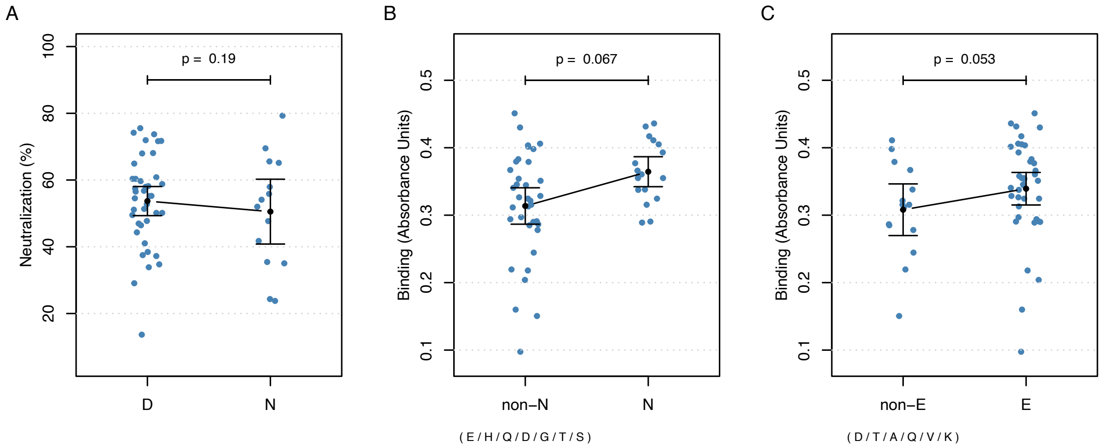


**Supplementary Figure 4: A) Association between site 653 and neutralization. B) Association between site 501 and binding. C) Association between site 533 and binding.** The black dots and lines indicate the mean and 95% confidence interval (CI) for each group. *P* values were calculated using linear regression model.

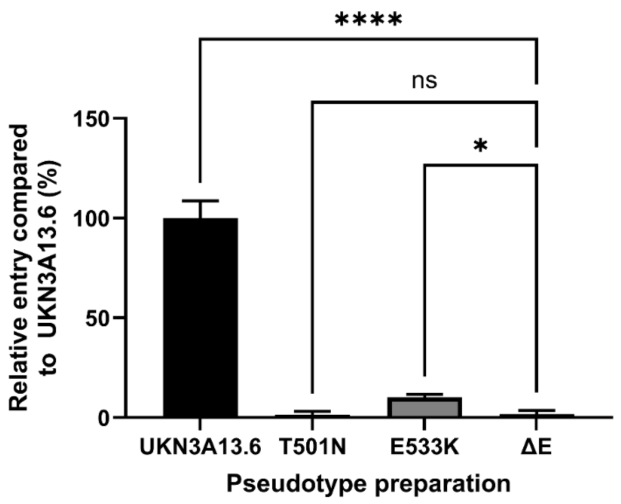


**Supplementary Figure 5: Infectivity of HCV mutants using E1/E2 Pseudo-particles.** Infection was performed using HuH7 cells, infected with pseudotypes bearing the wild-type UKN3A13.6 variant glycoprotein, or single aa mutants T501N or E533K. A preparation possessing pseudotypes created in the absence of E1E2 (ΔE) was used as a negative control. Statistical comparisons were performed using one-way ANOVA with Dunnett's correction for multiple comparisons. **** $p<0.0001$, * $p<0.05$, n.s. not significant.

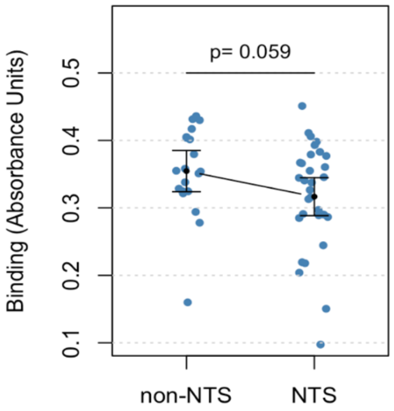


**Supplementary Figure 6: Association between glycosylation site N234 and binding.** The black dots and lines indicate the mean and 95% confidence interval (CI) for each group. *P* values were calculated using linear regression model. The non-NTS group comprised of NKS(7), NAS(6), NKT(2), NHS, NIS, NMS, NVS and NMS polymorphisms at this glycosylation site.

**Supplementary Table 6. The total count of potential N-linked glycosylation sites detected in each patient.** Most patients (48) exhibit 15 potential glycosylation sites, followed by a smaller number with 16, 13, or 14 sites. The data highlights variability in glycosylation site counts among patients.

| Total number of potential glycosylation sites | Patients |
|---|---|
| 13 | 2 |
| 14 | 1 |
| 15 | 48 |
| 16 | 3 |

**Supplementary Table 7. Association between the tested glycosylation motifs at 6 glycosylation sites and antibody response.** The table indicates the *p* values, effect size, standard error from the linear regression for each test of the association between the absence or presence of E1E2 N-linked glycosylation motifs and antibody binding and neutralization response. The names of the tested sites were relative to H77 polyprotein numbering. Associations at 20% FDR are highlighted in bold.

| Binding/ Neutralization | N-glycosites | Most associated Motif | p value | Effect size | Standard error | q value |
|---|---|---|---|---|---|---|
| **binding** | **N476** | **NIT** | **0.03499292** | **0.61327109** | **0.28207457** | **0.13997168** |
| **binding** | **N234** | **NTS** | **0.05858346** | **-0.5081009** | **0.26166753** | **0.1406003** |
| binding | N581 | NGS | 0.28424741 | -0.3369855 | 0.31047421 | 0.4167147 |
| binding | N533 | NET | 0.31253602 | 0.26895107 | 0.26332224 | 0.4167147 |
| binding | N430 | NDS | 0.82846376 | -0.0588983 | 0.27021831 | 0.90377865 |
| binding | N423 | NRT | 0.95930719 | -0.0139371 | 0.27166948 | 0.95930719 |
| **neutralization** | **N476** | **NIT** | **0.00435529** | **0.8938111** | **0.29765221** | **0.05226352** |
| neutralization | N234 | NTS | 0.02083243 | -0.6830836 | 0.28494859 | 0.12499457 |
| neutralization | N533 | NET | 0.05194934 | 0.57000856 | 0.28550219 | 0.1406003 |
| neutralization | N581 | NGS | 0.25163239 | 0.38400524 | 0.33012242 | 0.4167147 |
| neutralization | N423 | NRT | 0.30555152 | -0.3075108 | 0.29678391 | 0.4167147 |
| neutralization | N430 | NES | 0.63561672 | -0.1436828 | 0.30112701 | 0.76274006 |

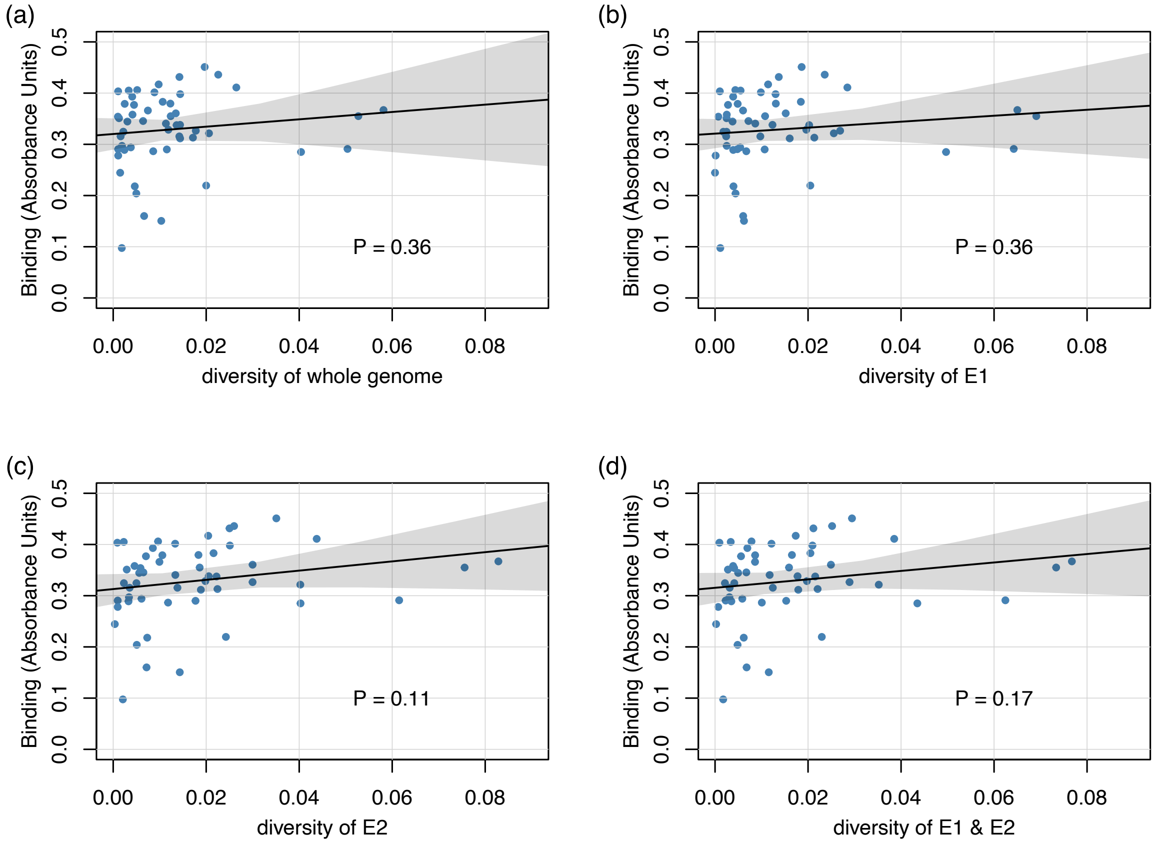


**Supplementary Figure 7A. Correlates of intra-patient viral nucleotide diversity and binding.** a) Correlation between diversity in whole viral sequences, b) E1 protein region, c) E2 protein region, d) E1 and E2 region together and baseline binding. The solid black lines show the best fit linear regression line and grey area indicates its 95% confidence interval. *P* values for the slopes are from the linear regression model.

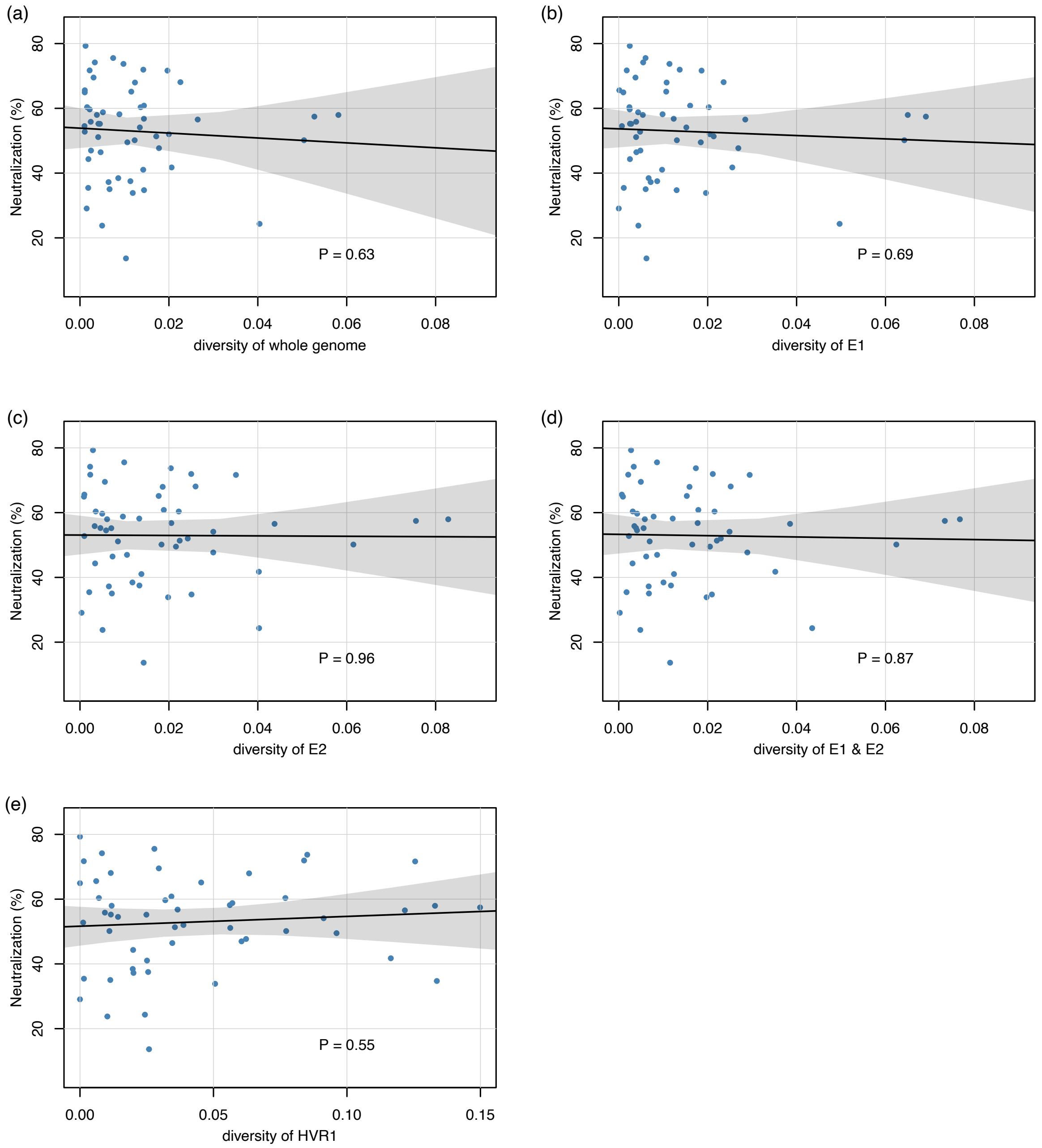


**Supplementary Figure 7B: Correlates of intra-patient viral nucleotide diversity and neutralization. a)** Correlation between diversity in whole viral sequences, **b)** E1 protein region, **c)** E2 protein region, **d)** E1 and E2 region together, **e)** HVR1 region and baseline binding. The solid black lines show the best fit linear regression line and grey area indicates its 95% confidence interval. *P* values for the slopes are from the linear regression model.

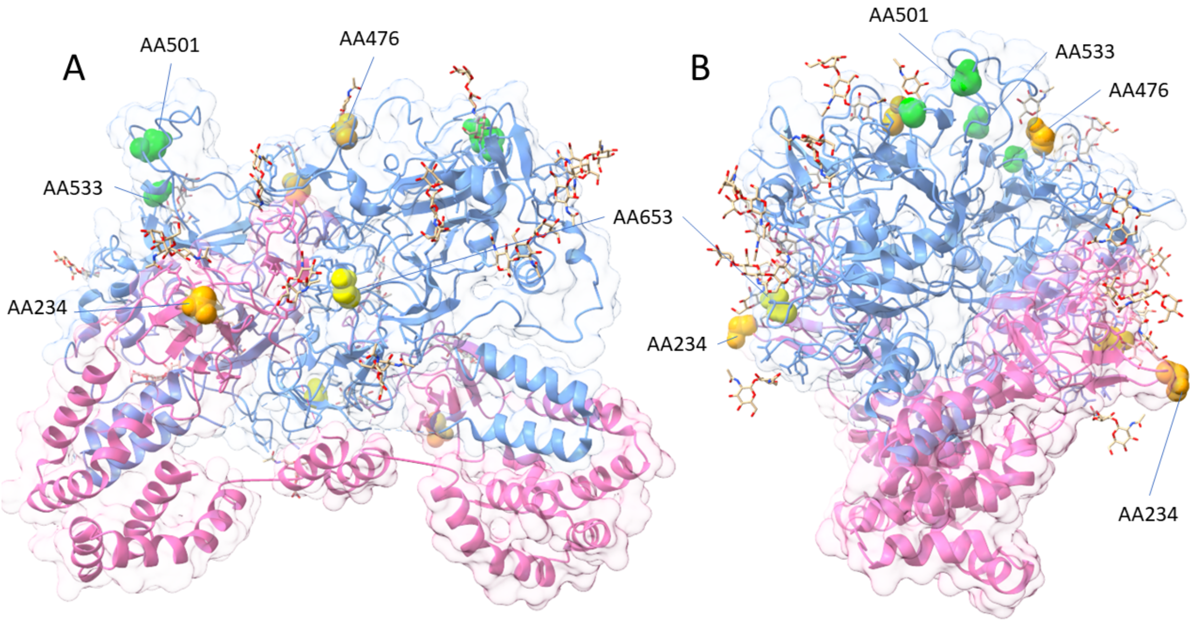


**Supplementary Figure 8. Location of sites associated with antibody neutralization/binding phenotype in a structure of the E1/E2 heterocomplex.** In this structure of E1/E2 (PDB 8RJJ), E1 is highlighted in pink, with E2 highlighted in blue. Carbohydrates associated with these proteins are presented as wireframes, with a space-filling representation of the amino acid side chains representing the two amino acids associated with neutralization phenotype (AA501 and AA533, green), and the amino acid associated with antibody binding (AA653, yellow). Asparagine residues 234 and 476 modified with *N*-linked glycans are highlighted in orange. The left figure is a 'side-on' presentation of the 'dimer of heterodimer' structure. The right one is for the structure rotated through 90$^{o}$ in a vertical axis.
